# Original research: Impact of surveillance colonoscopy on colorectal cancer incidence and mortality in Lynch syndrome - a national observational cohort study of patients in the English NHS 2010-2022

**DOI:** 10.64898/2026.04.16.26351020

**Authors:** Catherine Huntley, Lucy Loong, Corinne Mallinson, Tameera Rahman, Beth Torr, Sophie Allen, Isaac Allen, Hend Hassan, Yvonne Walburga Joko Fru, Daniela Tataru, Lizz Paley, Sally Vernon, Richard Houlston, David Muller, Fiona Lalloo, Adam Shaw, John Burn, Eva Morris, Marc Tischkowitz, Antonis C. Antoniou, Paul D.P. Pharoah, Kevin Monahan, Steven Hardy, Clare Turnbull

**Affiliations:** Division of Genetics and Epidemiology, The Institute of Cancer Research, Sutton, UK; National Disease Registration Service, NHS England, London, UK; Health Data Insight CIC, Cambridge, UK; Centre for Cancer Genetic Epidemiology, Department of Public Health and Primary Care, University of Cambridge, Cambridge, United Kingdom; School of Public Health, Faculty of Medicine, Imperial College London, London, UK; Manchester Centre for Genomic Medicine and North West Genomic Laboratory Hub, Manchester University NHS Foundation Trust, Manchester Academic Health Science Centre, Manchester, United Kingdom; Department of Genetics, Guy’s and St Thomas’ NHS Foundation Trust, London, UK; Translational and Clinical Research Institute, Newcastle University, Newcastle upon Tyne, UK; Applied Health Research Unit, Big Data Institute, Nuffield Department of Population Health, University of Oxford, Oxford, UK; Department of Genomic Medicine, Cambridge Biomedical Research Centre, National Institute for Health Research, University of Cambridge, Cambridge, United Kingdom; Department of Computational Biomedicine, Cedars-Sinai Medical Center, Los Angeles, CA, USA; The Lynch Syndrome and Family Cancer Clinic, St Mark’s Hospital and Academic Institute, Harrow, London, UK; Imperial College London, London, UK; Cancer Genetics Unit, The Royal Marsden NHS Foundation Trust, London, UK

## Abstract

**Background:** Lynch syndrome (LS) is a cancer susceptibility syndrome caused by germline pathogenic variants in DNA mismatch repair (MMR) genes. Due to increased risk of colorectal cancer (CRC), enhanced colonoscopic surveillance is recommended for heterozygote MMR-carriers.

**Objective:** Using a registry of English LS patients linked to digital National Health Service records, we aimed to assess adherence of MMR-carriers to national surveillance guidelines, and to determine the impact of surveillance on CRC incidence and mortality.

**Design:** We described the frequency of colonoscopies in 4,732 MMR-carriers and used logistic regression to determine predictors of surveillance adherence. For MMR-carriers with a record of surveillance and those without, we: estimated age-specific annual CRC incidence rates (AS-AIRs) and cumulative lifetime risks, assessed for stage-shift by comparing CRC stage distributions and stage-specific AS-AIRs, and estimated risks of death from CRC and any cause using Kaplan-Meier methods and Cox Proportional Hazards regression.

**Results:** Surveillance at a mean interval of ≤ 3 years (n=3028) was associated with a decrease in CRC-specific and all-cause mortality, without an associated change in total CRC incidence, even after multivariate adjustment. No strong evidence of stage-shift was observed. Colonoscopic surveillance at a mean interval of ≤ 2 years (n=1569) was associated with an increase in total CRC incidence. Incidence of early-stage cancers was also higher, with no corresponding decrease in late-stage cancers, which may reflect the short follow-up period or the impact of overdiagnosis.

**Conclusion:** The observed reduction in all-cause mortality amongst regularly-surveilled MMR-carriers may indicate an impact of surveillance on CRC-specific mortality, though in the context of a non-randomised study likely reflects the influence of selection bias.

**KEY MESSAGES OF ARTICLE:** *What is already known on this topic:* Regular surveillance colonoscopy is recommended in Lynch syndrome, though evidence to support this remains mixed. We searched PubMed for articles published from inception to 01/05/2024 using the terms “Lynch syndrome”, “HNPCC”, “colonoscopy”, “sigmoidoscopy”, “surveillance”, and “screening”. We found one controlled trial and several small analytical studies dating from the early 2000s which compared surveilled and non-surveilled populations and found surveillance to be associated with reduced colorectal cancer (CRC) incidence and improved survival. More recent longitudinal observational studies, most without comparator groups, found a high incidence of CRC in LS populations despite being resident in countries where surveillance was recommended. A small number of studies directly assessed time since last colonoscopy against CRC incidence and stage with mixed findings. Finally, cross-sectional comparisons between countries of CRC incidence rates and surveillance interval recommendations found no relationship between the two^1,2^.

*What this study adds:* Here, we conduct an observational cohort study on a large national cohort of MMR germline pathogenic variant (GPV) carriers (MMR-carriers) in England (n=4,732), comparing CRC incidence and mortality in individuals with a record of regular surveillance to those without. Through linkage of the English National Lynch Syndrome Registry to Hospital Episodes Statistics data, we are uniquely able to study a comprehensive national population of MMR-carriers and identify the dates on which colonoscopies were undertaken over time, allowing assessment of adherence to national surveillance guidelines and the impact this has on CRC outcomes. Notably, receipt of regular colonoscopy was strongly associated with deprivation as well as ethnicity. The results show that regular surveillance at an average interval of 3 years (or less) is not associated with a reduction in CRC incidence when compared to less frequent surveillance, but an apparent decrease in both CRC-specific and overall mortality is observed, even after adjustment for confounding variables. Conversely, regular surveillance at an average interval of 2 years (or less) is associated with an increase in CRC incidence when compared to less frequent surveillance, which may suggest increased diagnosis of early-stage cancers or, due to the absence of a reduction in late-stage cancers, overdiagnosis. The observed impact of surveillance on overall mortality may demonstrate the impact of surveillance on CRC-specific mortality, or, in the context of an observational (non-randomised) study, indicate that the results are subject to selection bias.

*How this study might affect research, practice, or policy:* Evidence for the benefit of surveillance colonoscopy remains mixed. Whilst polypectomy would be anticipated to prevent CRC development (thus reducing CRC incidence), several studies have observed increased frequency of CRCs in MMR-carriers undergoing frequent surveillance colonoscopy, which may reflect overdiagnosis. The selection bias inherent to observational studies of surveillance renders mortality outcomes challenging to interpret. Randomised controlled trials of colonoscopic surveillance in MMR-carriers are required for effectiveness of this intervention to be accurately assessed. Given ethical and feasibility challenges, randomised controlled trials might be complemented by quasi-experimental designs using advanced observational methods for assessing effectiveness.

## INTRODUCTION

Lynch syndrome (LS) is a cancer susceptibility syndrome caused by germline pathogenic variants (GPVs) in DNA mismatch repair (MMR) genes (*MLH1*, *MSH2*, *MSH6*, and *PMS2*). Heterozygote carriers of MMR gene GPVs (MMR-carriers) have a high lifetime risk of colorectal cancer (CRC) compared to the general population, and thus may be offered additional preventative interventions, including regular surveillance colonoscopy with removal of polyps (adenomas). Surveillance in MMR-carriers is theorised to prevent CRCs through disruption of the adenoma-carcinoma sequence, thus reducing CRC incidence, and to facilitate earlier detection, thus reducing CRC mortality.

In the UK, Europe, and elsewhere, enhanced endoscopic surveillance has been recommended for MMR-carriers since the early 2000s. In 2010, the British Society of Gastroenterology (BSG), Association of Coloproctology of Great Britain and Ireland (ACPGBI), and United Kingdom Cancer Genetics Group (UKCGG) updated their surveillance guidelines to specify 18-monthly colonoscopies from the age of 25-75 for all MMR-carriers, and again in 2020 to specify two-yearly colonoscopies from age 25 for *MLH1-*and *MSH2*-carriers, and from age 35 for *MSH6*-and *PMS2*-carriers^1,2^.

These recommendations were introduced following the publication of, to this day, the only (non-randomised) controlled trial of colonoscopic surveillance in LS, in which a reduction in CRC incidence and mortality was observed in LS individuals receiving regular surveillance compared to those who were not^3,4^. These results were replicated in several small (likewise non-randomised) studies, which found a reduction in CRC incidence and mortality in surveilled MMR-carriers^5–7^. In the 2010s, the advent of large multi-country prospective registries of MMR-carriers, such as the Prospective Lynch Syndrome Database (PLSD), ushered in a new era of international longitudinal studies of LS populations, which demonstrated MMR-carriers to have persistently high CRC incidence rates despite regular surveillance^8–10^. Comparisons of LS populations across countries with differing surveillance intervals report no reduction in CRC incidence with surveillance at an interval of 1-2 years compared to 3 years^11,12^. Yet other studies have shown there both to be an association between surveillance interval and CRC development, and conversely, no association between surveillance interval and CRC stage^12–16^.

The evidence for the impact on incidence and mortality of regular colonoscopic surveillance thus remains mixed. A series of explanations have been proffered for the continued high incidence of CRC in MMR-carriers resident in countries where regular surveillance is recommended, including a rapid adenoma-carcinoma sequence, alternative pathways of tumour development, overdiagnosis, and low-quality surveillance^17^. However, without direct assessment of adherence, and analysis of a relevant comparator group, the possibility of an even higher CRC incidence in non-surveilled populations of MMR-carriers cannot be excluded. Thus, the generation of robust evidence to underpin surveillance recommendations has been hampered by a lack of large, comprehensive MMR-carrier cohorts in which the adherence of participants to regular surveillance programmes is known.

The English National Lynch Syndrome Registry (ENLSR), a nationally comprehensive register of MMR-carriers in England, affords an opportunity to address some of these challenges by providing a large cohort of LS individuals whose participation in surveillance can be determined through linkage to Hospital Episode Statistics (HES) data^18^. In this analysis, we aimed to use the ENLSR cohort to assess adherence of MMR-carriers to national surveillance guidelines, and to determine the impact of regular surveillance on CRC incidence and mortality.

## METHODS

### Study design and participants

This was an observational cohort study. Data originated from the English National Lynch Syndrome Registry (ENLSR)^18^, Germline Genetic Testing Dataset (GGT)^19^, National Cancer Registration Dataset (NCRD)^20^, and Hospital Episode Statistics Admitted Patient Care (HES APC)^21^ and HES Outpatients (HES OP) datasets. A cohort for analysis was constructed from an extract of the ENLSR censored at the 28^th^ of November 2024, comprising the total known LS population in England on that date. All LS-carriers included in the registry have a clinical diagnosis of LS and are known carriers of a GPV in an MMR gene. Demographic (date of birth, person stated gender [hereafter referred to as sex], NHS number, ethnicity, postcode of residence, vital status, date of death) and genetic (MMR gene, date of clinical LS diagnosis) data were drawn from the ENLSR. Dates of molecular diagnosis for individuals (without a clinical date of LS diagnosis) were drawn from the GGT. Cancer (date of diagnosis, type of cancer, stage of cancer at diagnosis), and additional demographic (ethnicity, underlying cause of death) data were obtained from the NCRD. Colonoscopy (type of procedure, date of procedure) and further demographic (ethnicity, Index of Multiple Deprivation [IMD] quintile [of residence]) data were obtained from HES. ENLSR data were cleaned and de-duplicated using methods previously described^18^. Datasets were linked using unique patient identifiers. Inclusion criteria were: i) known demographic details (NHS number, date of birth, sex), ii) known genetic details (MMR gene and date of LS diagnosis), iii) diagnosed with LS prior to any colorectal cancer (date of LS diagnosis must precede date of first colorectal diagnosis by at least 60 days, no individuals with prior CRC are included), and iv) eligible for surveillance colonoscopy (according to 2020 guidelines) during the study period (1^st^ of January 2010 and 31^st^ of December 2022). Further details on data sources, linkage, and cleaning are provided in the supplementary methods.

### Colonoscopy analyses

The outcome of interest was ‘*3-year-adherent’*, defined as a mean surveillance interval of ≤ 3 years during the follow up period. The follow up period was defined as the time for which each individual was eligible for surveillance, beginning at the latest of: study start (01/01/2010), date of (clinical or molecular) LS diagnosis, or individual eligibility for surveillance (aged 25 for *MLH1*/*MSH2*-carriers, 35 for *MSH6*/*PMS2*-carriers); and ending at the earliest of: study end (31/12/2022), date of first CRC diagnosis, date of death/embarkation, or date individual eligibility for surveillance ends (age 75). Mean surveillance interval (MSVI) was calculated for each individual by dividing the total follow up time by the number of surveillance colonoscopies during that period. Surveillance colonoscopies were identified by a list of pre-defined OPCS-4 codes (Supplementary Table 1), grouped into single events if occurring within 90 days of one another, and assigned the date of the first procedure. Logistic regression was used to model the relationship between explanatory variables (MMR gene, sex, age at start of follow up period, ethnicity, and IMD quintile) and the binary outcome variable ‘*3-year-adherent’*. Univariable models were constructed for each explanatory variable in turn, and associated variables carried through to the multivariable model. p-values for linear trend were calculated for ordinal variables using linear polynomial contrasts.

### Colorectal cancer incidence analyses

The outcome of interest was a first diagnosis of invasive colorectal cancer (ICD-10 codes C18, C19, C20), and the explanatory variable was *‘3-year-adherent’* (MSVI ≤ 3 years). The follow up period was the same as for the colonoscopy analyses. Age-specific annual incidence rates (AS-AIRs) per 100,000 person years for five-year age bands were calculated in i) all MMR-carriers combined, and ii) MMR-carriers of each individual gene, for ‘*3-year-adherent’* MMR-carriers compared to non *‘3-year-adherent’* MMR-carriers. The z-test was used to test for a statistically significant difference between analysis groups. Life table methods were used to estimate the cumulative risk of CRC from age 25-74 for the same analysis groups. For all MMR-carriers combined, the possibility of stage-shift between analysis groups (‘*3-year-adherent*’ vs non ‘*3-year-adherent*’) was investigated by comparing stage distributions using the chi-square test, and by calculating stage-and age-specific annual incidence rates in five year age bands for early-stage cancers (stages 1 and 2) and late-stage cancers (stages 3 and 4). The analysis was then repeated using ‘*2-year-adherent’* (MSVI ≤ 2 years) as the explanatory variable to examine the impact of more frequent colonoscopies.

### Mortality analyses

The follow up period for the mortality analysis began at the latest of: the study start date (01/01/2010), the date of (clinical or molecular) LS diagnosis, or the date the individual became eligible for surveillance (aged 25 for *MLH1*/*MSH2*-carriers or 35 for *MSH6*/*PMS2*-carriers); and ended at the earliest of: date of death, date of embarkation, or study end date. The outcomes of interest were a) death from any cause during the study period, and b) death from CRC during the study period. CRC-specific death was defined as any death where a CRC ICD-10 code (C18, C19, C20) appeared as a cause code in any section (1A, 1B, 1C, or 2) of the death certificate. Kaplan-Meier methods were used to estimate the risk of death from i) CRC-only, ii) any cause, and iii) any non-CRC cause for individuals with an MSVI ≤ 3 years (*‘3-year-adherent’*) compared to those with an MSVI > 3 years (non *‘3-year-adherent’*). Cox proportional hazards modelling was used to estimate hazard ratios (HRs) for associations between MSVI ≤ 3 years (*‘3-year-adherent’*), MMR gene, sex, age at start of follow up period, IMD quintile, and risk of death from i) CRC only and ii) any cause. For each variable, the proportional hazards assumption was tested using Schoenfeld residuals, and only variables meeting this assumption were included in the model.

### Sensitivity analyses

To investigate the potential for skewing of the MSVI in individuals with short follow up periods, a second cohort was constructed excluding all individuals from the primary cohort who had less than one year of follow up. The CRC incidence analysis protocol described above was repeated.

### Patient and public involvement

This work uses data that has been provided by patients and collected by the NHS as part of their care and support. The data are collated, maintained and quality assured by the National Disease Registration Service, which is part of NHS England. NDRS is committed to Patient and Public Involvement (PPI), including providing publicly downloadable reports, running public awareness campaigns and webinars, and providing opportunities for public consultation and representation^22^.

## RESULTS

### Cohort description

Following exclusions, the cohort consisted of 4,732 MMR-carriers (Supplementary Figure 1). The cohort was majority white (n=3,908, 82.6%) and female (n=2,908, 61.5%), with representation from all ten-year age bands from age 25-74, and all IMD quintiles. The MMR genes in which GPVs were most commonly identified were *MSH2* (n=1,856 [including 25 individuals with *EPCAM* deletions], 39.2%) and *MLH1* (n=1,315, 27.8%), followed by *MSH6* (n=1,011, 21.4%) and *PMS2* (n=550, 11.6%). Further details are provided in Table 1.

**Table 1.** Description of Lynch syndrome cohort and analysis groups.

|  |  | Total |  | MSVI 2 years |  |  |  | MSVI 3 years |  |  |  |
| --- | --- | --- | --- | --- | --- | --- | --- | --- | --- | --- | --- |
|  |  |  |  | ≤ 2 years |  | > 2 years |  | ≤ 3 years |  | > 3 years |  |
|  |  | n | % | n | % | n | % | n | % | n | % |
| **Total** |  | 4732 |  | 1569 | 33.2 | 3163 | 66.8 | 3028 | 64.0 | 1704 | 36.0 |
| **MMR Gene** | *MLH1* | 1315 | 27.8 | 410 | 26.1 | 905 | 28.6 | 837 | 27.6 | 478 | 28.1 |
|  | *MSH2* | 1856 | 39.2 | 608 | 38.8 | 1248 | 39.5 | 1224 | 40.4 | 632 | 37.1 |
|  | *MSH6* | 1011 | 21.4 | 371 | 23.6 | 640 | 20.2 | 655 | 21.6 | 356 | 20.9 |
|  | *PMS2* | 550 | 11.6 | 180 | 11.5 | 370 | 11.7 | 312 | 10.3 | 238 | 14.0 |
|  | Unknown | 0 | 0.0 | 0 | 0.0 | 0 | 0.0 | 0 | 0.0 | 0 | 0.0 |
| **Sex** | Male | 1824 | 38.5 | 629 | 40.1 | 1195 | 37.8 | 1190 | 39.3 | 634 | 37.2 |
|  | Female | 2908 | 61.5 | 940 | 59.9 | 1968 | 62.2 | 1838 | 60.7 | 1070 | 62.8 |
|  | Unknown | 0 | 0.0 | 0 | 0.0 | 0 | 0.0 | 0 | 0.0 | 0 | 0.0 |
| **Age at start of follow up period** | 25-34 | 1208 | 25.5 | 363 | 23.1 | 845 | 26.7 | 805 | 26.6 | 403 | 23.7 |
|  | 35-44 | 1209 | 25.5 | 398 | 25.4 | 811 | 25.6 | 781 | 25.8 | 428 | 25.1 |
|  | 45-54 | 1034 | 21.9 | 376 | 24.0 | 658 | 20.8 | 702 | 23.2 | 332 | 19.5 |
|  | 55-64 | 869 | 18.4 | 296 | 18.9 | 573 | 18.1 | 511 | 16.9 | 358 | 21.0 |
|  | 65-74 | 412 | 8.7 | 136 | 8.7 | 276 | 8.7 | 229 | 7.6 | 183 | 10.7 |
|  | Unknown | 0 | 0.0 | 0 | 0.0 | 0 | 0.0 | 0 | 0.0 | 0 | 0.0 |
| **Ethnicity** | White | 3908 | 82.6 | 1369 | 87.3 | 2539 | 80.3 | 2651 | 87.5 | 1257 | 73.8 |
|  | Asian | 207 | 4.4 | 58 | 3.7 | 149 | 4.7 | 108 | 3.6 | 99 | 5.8 |
|  | Black | 30 | 0.6 | 8 | 0.5 | 22 | 0.7 | 15 | 0.5 | 15 | 0.9 |
|  | Mixed | 31 | 0.7 | 10 | 0.6 | 21 | 0.7 | 19 | 0.6 | 12 | 0.7 |
|  | Other | 98 | 2.1 | 30 | 1.9 | 68 | 2.1 | 57 | 1.9 | 41 | 2.4 |
|  | Unknown | 458 | 9.7 | 94 | 6.0 | 364 | 11.5 | 178 | 5.9 | 280 | 16.4 |
| **IMD Quintile** | Q 1 - most deprived | 727 | 15.4 | 240 | 15.3 | 487 | 15.4 | 452 | 14.9 | 275 | 16.1 |
|  | Q 2 | 844 | 17.8 | 288 | 18.4 | 556 | 17.6 | 536 | 17.7 | 308 | 18.1 |
|  | Q 3 | 939 | 19.8 | 283 | 18.0 | 656 | 20.7 | 572 | 18.9 | 367 | 21.5 |
|  | Q 4 | 1031 | 21.8 | 355 | 22.6 | 676 | 21.4 | 688 | 22.7 | 343 | 20.1 |
|  | Q 5 - least deprived | 1042 | 22.0 | 363 | 23.1 | 679 | 21.5 | 702 | 23.2 | 340 | 20.0 |
|  | Unknown | 149 | 3.1 | 40 | 2.5 | 109 | 3.4 | 78 | 2.6 | 71 | 4.2 |

Numbers (n) and percentages (%) are provided for the entire cohort (Total), and analysis groups. Carriers for EPCAM deletions (n=25) are included in the *MSH2* category. Sex is person stated gender. Ethnicity is self-described. IMD quintile is determined by LSOA of residence. MSVI, Mean surveillance interval.

### Colonoscopy adherence

Across all 4,732 MMR-carriers, there was a total of 27,866 years of follow up for the colonoscopy analyses (median 4.88) (Supplementary Table 2). 3,788 MMR-carriers (80.1%) had a record of at least one colonoscopy in HES. *MSH2*-carriers were most likely to have a record of at least one colonoscopy at 83.0% (n=1,541) of individuals, followed by *MLH1*-carriers (81.7%, n=1,074), *MSH6*-carriers (78.5%, n=794), and *PMS2*-carriers (68.9%, n=379) (Supplementary Table 3). The mean colonoscopies per person for the whole cohort was 2.37, ranging from 1.46 for *PMS2*-carriers, to 2.68 for *MSH2*-carriers. By gene, the MSVI for all MMR-carriers combined was 2.51, and 2.58, 2.61, 2.36, and 2.22 for *MLH1*, *MSH2*, *MSH6*, and *PMS2* carriers, respectively.

3,788 first colonoscopies were conducted in the cohort, of which 65.8% (n=2,492) took place within 12 months of the date of eligibility for colonoscopic surveillance. The median time to first colonoscopy from the date of eligibility was 214.5 days. 7,413 ‘subsequent’ colonoscopies were conducted in the cohort, of which 91.2% (n=6,760) took place within 36 months of the previous colonoscopy, and 41.2% within 24 months (Supplementary Table 3). To characterise an individual’s adherence to surveillance guidelines throughout the course of the study period, we applied a pragmatically permissive definition, reasoning that while colonoscopies were recommended at 18-24 month intervals throughout the study period, delays may have occurred. For this reason, we considered individuals to be 3-year-adherent where their mean surveillance interval (MSVI) is ≤ 3 years across their duration of follow-up.

Univariable logistic regression analyses of persons showed MMR gene, age at start of follow up period, ethnicity, and IMD quintile to be statistically significantly associated with likelihood of being ‘*3-year-adherent’* (MSVI ≤ 3 years), while sex was not. In multivariable logistic regression, including adjustment for all variables, age at start of follow up was significantly associated with being ‘*3-year-adherent’*, as well as IMD quintile. The likelihood of being *‘3-year-adherent’* decreased as age increased (p-trend = 0.002) and increased as IMD quintile moved from most to least deprived (p-trend = 0.001) (Table 2).

**Table 2.**
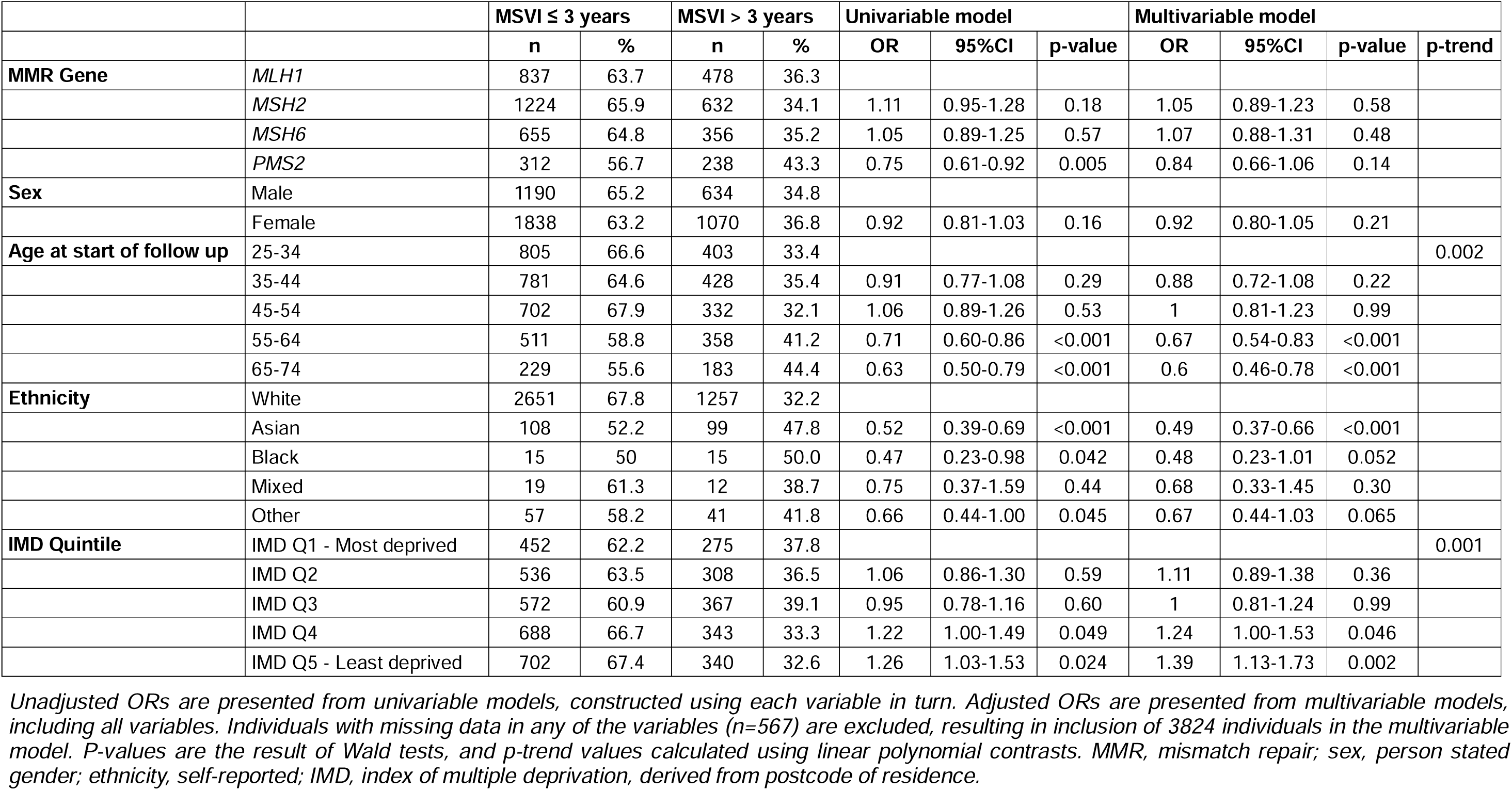
Uni-and multi-variable logistic regression models for ‘*3-year-adherence’* (mean surveillance interval ≤ 3 years)

| | | MSVI $\leq 3$ years | | MSVI $> 3$ years | | Univariable model | | | Multivariable model | | | |
| --- | --- | --- | --- | --- | --- | --- | --- | --- | --- | --- | --- | --- |
|  |  | n | % | n | % | OR | 95%CI | p-value | OR | 95%CI | p-value | p-trend |
| **MMR Gene** | *MLH1* | 837 | 63.7 | 478 | 36.3 |  |  |  |  |  |  |  |
|  | *MSH2* | 1224 | 65.9 | 632 | 34.1 | 1.11 | 0.95-1.28 | 0.18 | 1.05 | 0.89-1.23 | 0.58 |  |
|  | *MSH6* | 655 | 64.8 | 356 | 35.2 | 1.05 | 0.89-1.25 | 0.57 | 1.07 | 0.88-1.31 | 0.48 |  |
|  | *PMS2* | 312 | 56.7 | 238 | 43.3 | 0.75 | 0.61-0.92 | 0.005 | 0.84 | 0.66-1.06 | 0.14 |  |
| **Sex** | Male | 1190 | 65.2 | 634 | 34.8 |  |  |  |  |  |  |  |
|  | Female | 1838 | 63.2 | 1070 | 36.8 | 0.92 | 0.81-1.03 | 0.16 | 0.92 | 0.80-1.05 | 0.21 |  |
| **Age at start of follow up** | 25-34 | 805 | 66.6 | 403 | 33.4 |  |  |  |  |  |  | 0.002 |
|  | 35-44 | 781 | 64.6 | 428 | 35.4 | 0.91 | 0.77-1.08 | 0.29 | 0.88 | 0.72-1.08 | 0.22 |  |
|  | 45-54 | 702 | 67.9 | 332 | 32.1 | 1.06 | 0.89-1.26 | 0.53 | 1 | 0.81-1.23 | 0.99 |  |
|  | 55-64 | 511 | 58.8 | 358 | 41.2 | 0.71 | 0.60-0.86 | <0.001 | 0.67 | 0.54-0.83 | <0.001 |  |
|  | 65-74 | 229 | 55.6 | 183 | 44.4 | 0.63 | 0.50-0.79 | <0.001 | 0.6 | 0.46-0.78 | <0.001 |  |
| **Ethnicity** | White | 2651 | 67.8 | 1257 | 32.2 |  |  |  |  |  |  |  |
|  | Asian | 108 | 52.2 | 99 | 47.8 | 0.52 | 0.39-0.69 | <0.001 | 0.49 | 0.37-0.66 | <0.001 |  |
|  | Black | 15 | 50 | 15 | 50.0 | 0.47 | 0.23-0.98 | 0.042 | 0.48 | 0.23-1.01 | 0.052 |  |
|  | Mixed | 19 | 61.3 | 12 | 38.7 | 0.75 | 0.37-1.59 | 0.44 | 0.68 | 0.33-1.45 | 0.30 |  |
|  | Other | 57 | 58.2 | 41 | 41.8 | 0.66 | 0.44-1.00 | 0.045 | 0.67 | 0.44-1.03 | 0.065 |  |
| **IMD Quintile** | IMD Q1 - Most deprived | 452 | 62.2 | 275 | 37.8 |  |  |  |  |  |  | 0.001 |
|  | IMD Q2 | 536 | 63.5 | 308 | 36.5 | 1.06 | 0.86-1.30 | 0.59 | 1.11 | 0.89-1.38 | 0.36 |  |
|  | IMD Q3 | 572 | 60.9 | 367 | 39.1 | 0.95 | 0.78-1.16 | 0.60 | 1 | 0.81-1.24 | 0.99 |  |
|  | IMD Q4 | 688 | 66.7 | 343 | 33.3 | 1.22 | 1.00-1.49 | 0.049 | 1.24 | 1.00-1.53 | 0.046 |  |
|  | IMD Q5 - Least deprived | 702 | 67.4 | 340 | 32.6 | 1.26 | 1.03-1.53 | 0.024 | 1.39 | 1.13-1.73 | 0.002 |  |

*Unadjusted ORs are presented from univariable models, constructed using each variable in turn. Adjusted ORs are presented from multivariable models, including all variables. Individuals with missing data in any of the variables (n=567) are excluded, resulting in inclusion of 3824 individuals in the multivariable model. P-values are the result of Wald tests, and p-trend values calculated using linear polynomial contrasts. MMR, mismatch repair; sex, person stated gender; ethnicity, self-reported; IMD, index of multiple deprivation, derived from postcode of residence.*

### Incidence of colorectal cancer

334 CRCs were diagnosed in the cohort during the follow up period: 138 in *MLH1*-carriers, 167 in *MSH2*-carriers, 23 in MSH6-carriers, and 6 in PMS2-carriers (Supplementary Table 4). This low incidence likely reflects the relatively short follow-up period available. 7.7% (n=233) of MMR-carriers who were ‘*3-year-adherent*’ were diagnosed with a CRC, compared to 5.9% (n=101) of those who were not ‘*3-year-adherent’* (Supplementary Table 5). Examining individual MMR genes and then all genes combined, comparing MMR-carriers who were ‘*3-year-adherent’* and those who were not, there was no statistically significant difference in the five-year age-specific annual incidence rates (AS-AIRs) of CRC, nor in the cumulative risk of CRC ( **Error! Reference source not found.**, Supplementary Tables 6 and 7). There was evidence of a change in the stage distribution of CRCs at diagnosis in *MLH*1-carriers who were *‘3-year-adherent’* compared to those who were not (p=0.024), with a higher proportion (and absolute number) of stage 1 cancers (Supplementary Table 5). There was no evidence for significant change in the stage distribution for other MMR genes. Considering all MMR-carriers combined, there was no significant difference between the *‘3-year-adherent’* group compared to the non ‘*3-year-adherent’* for the AS-AIRs for either early stage (stage 1 and 2 combined) or late stage (stage 3 and 4) CRCs (**Error! Reference source not found.**, Supplementary Table 8).

Having used a pragmatically permissive definition adherence of MSVI ≤ 3 years, we next sought to examine how CRC incidence was influenced by a more frequent colonoscopy regimen. We repeated the analysis, considering individuals as adherent if their MSVI was ≤ 2 years (‘*2-year-adherent’*). The differences in CRC incidence between analysis groups were more marked: 10.0% (n=157) of MMR-carriers who were ‘*2-year-adherent*’ were diagnosed with a CRC, compared to 5.6% (n=177) of those who were not (Supplementary Table 9). AS-AIR point estimates were higher in *‘2-year-adherent’* MMR-carriers for all age groups, with a statistically significant difference for ages 30-34, 40-44, 50-54, and 55-59 (Supplementary Table 10). The cumulative risk of CRC diagnosis was higher at all ages in the *‘2-year-adherent’* group for all MMR-carriers combined, with the difference in risk increasing with age. At age 50, the cumulative risk of CRC in the *‘2-year-adherent’* group was 33.4% (95%CI 25.5-41.4%) and 14.1% (95%CI 10.7-17.4%) in ‘*non-2-year-adherent’* group, rising to 63.4% (95%CI 53.3-73.5%) and 37.4% (95%CI 31.8-42.9%) by age 74, respectively (Supplementary Table 11). A similar pattern was present for each MMR gene in isolation (**Error! Reference source not found.Error! Reference source not found.**). There was evidence of a change in the stage distribution of CRCs between the ‘*2-year-adherent’* and non ‘*2-year-adherent’* groups for all genes combined (p=0.040) and MLH1-carriers only (p=0.047) (Supplementary Table 9). AS-AIRS for stage 1 and 2 cancers trended higher across all age groups in ‘*2-year-adherent*’ MMR-carriers, with a statistically significant increase in early-stage cancers in *‘2-year-adherent’* 30–34-year-olds and 55–59-year-olds (p<0.05). By contrast, there was no evidence of a statistically significant difference in the AS-AIRs for late-stage cancers between the *‘2-year-adherent’* and *‘2-year-non-adherent’* groups in any age bands (**Error! Reference source not found.**, Supplementary Table 12).

### Mortality analyses

There were 5 CRC-specific deaths amongst 3,028 *‘3-year-adherent’* MMR-carriers compared to 17 amongst 1,704 non-*’3-year-adherent’* MMR-carriers. The cumulative risk of CRC-specific death at 5 and 10 years was 0.11% (95CI 0.00-0.24%) and 0.29% (95%CI 0.01-0.58%), respectively, in *‘3-year-adherent’* MMR-carriers, and 0.61% (95%CI 0.15-1.07%) and 1.57% (95%CI 0.67-2.47%) in non-*’3-year-adherent’* MMR-carriers (p<0.001) (Supplementary Table 13, Figure 3). Following adjustment for sex, age at follow up start, and IMD quintile, an MSVI ≤ 3 years was associated with a reduction in risk of CRC-specific death (HR 0.14, 95%CI 0.05-0.39) (Supplementary Table 14).

**Figure 1.**
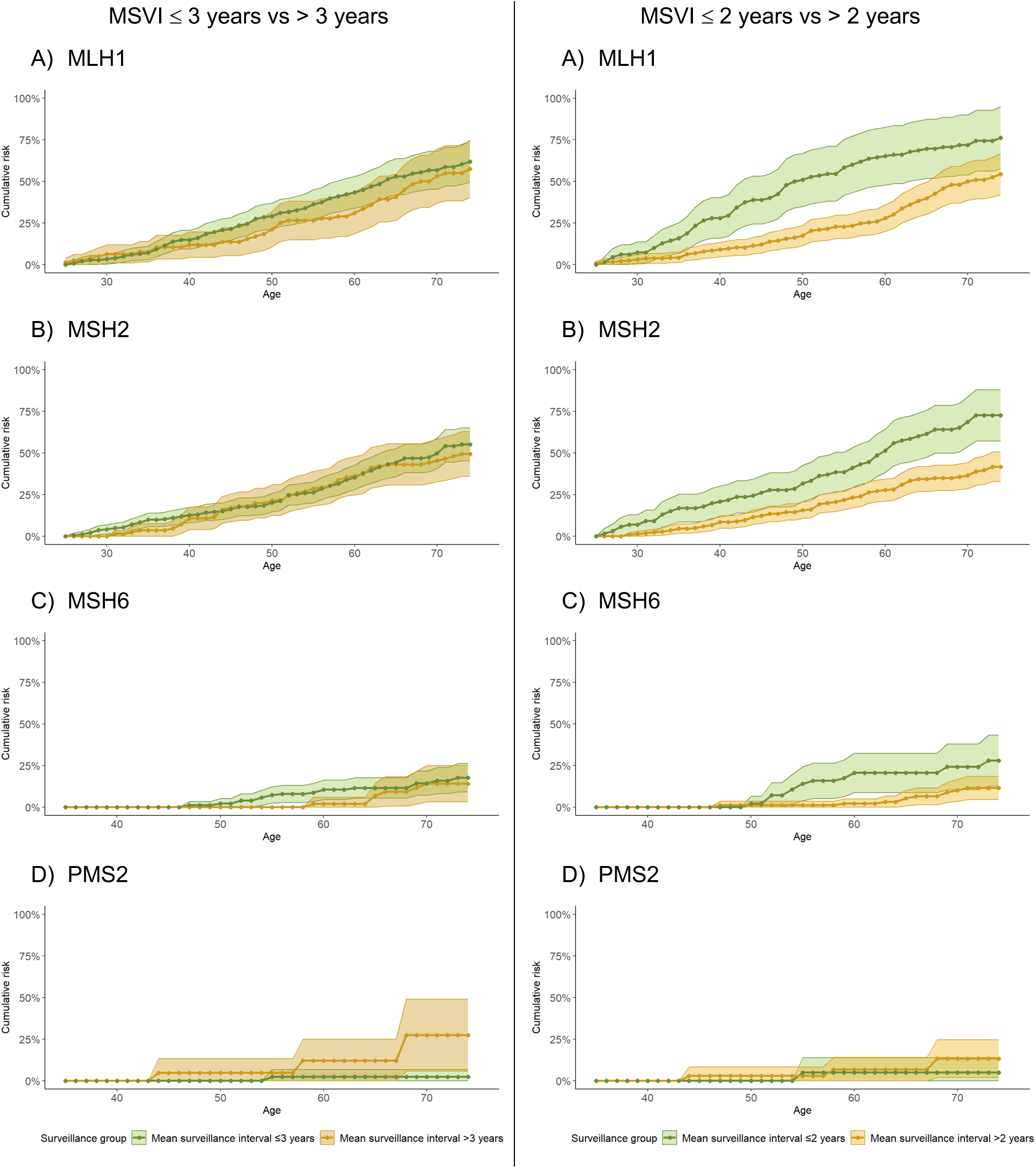
Cumulative risk (in %) of colorectal cancer in A) MLH1, B) MSH2, C) MSH6, and D) PMS2 MMR pathogenic variant carriers by mean surveillance interval: left panel. ≥ **3 years vs > 3 years, right panel** ≥ **2 years vs > 2 years** Solid points represent estimates of the cumulative risk, calculated using the life table method. Shaded bands represent the 95% confidence interval, estimated from the standard error derived using Greenwood’s formula. Assumed zero risk below the age of 25 for MLH1/MSH2-carriers, and below the age of 35 for MSH6/PMS2-carriers.

**Figure 2.**
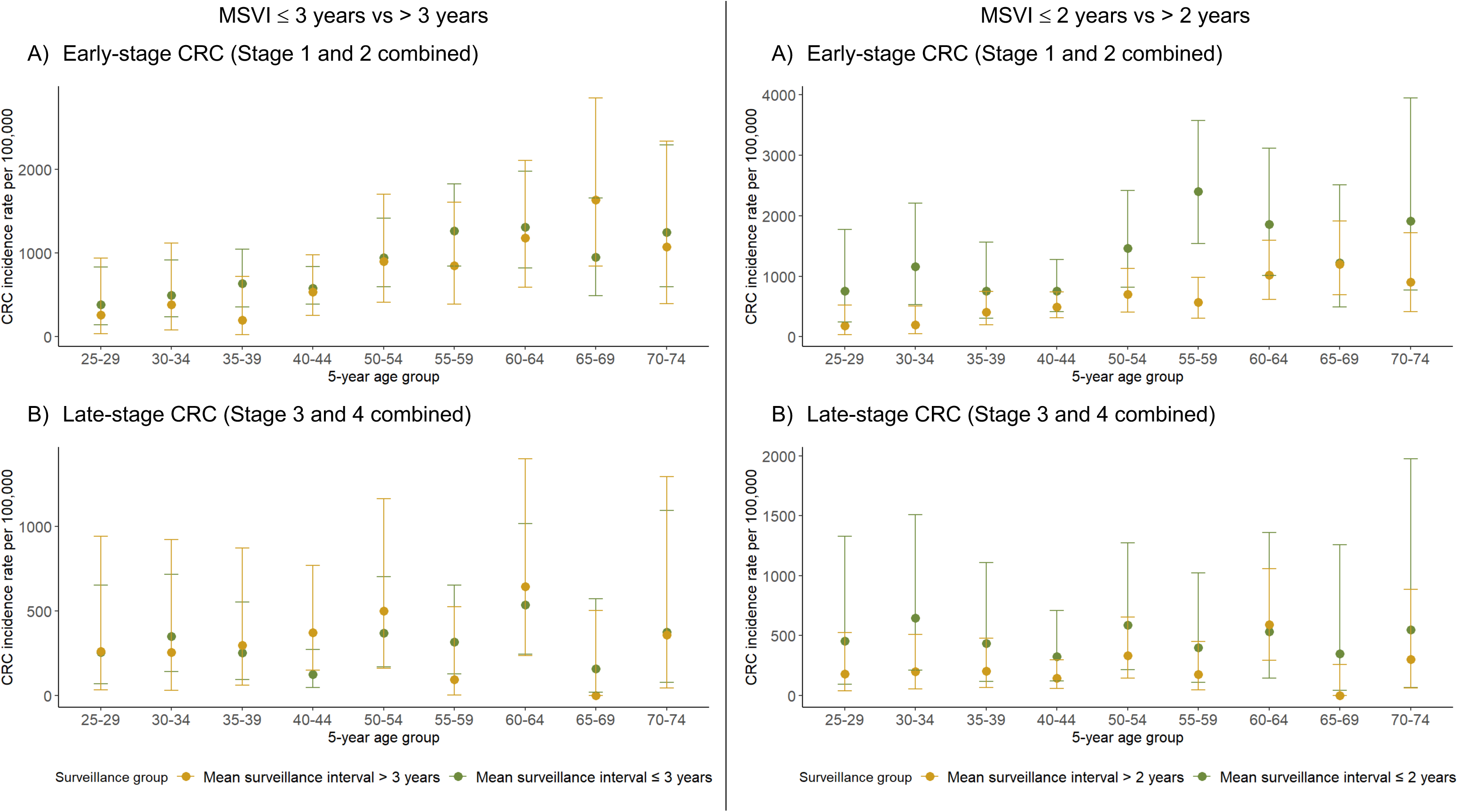
Stage-and age-specific five-year annual incidence rates for A) Early stage (Stage 1 and 2) colorectal cancers and B) Late stage (Stage 3 and 4) colorectal cancers by mean surveillance interval: left panel. ≥ **3 years vs > 3 years, right panel** ≥ **2 years vs > 2 years** Only first cancers are included. Tumours are staged at diagnosis. 95% confidence intervals are calculated using Byar’s method. CRC, colorectal cancer.

**Figure 3.**
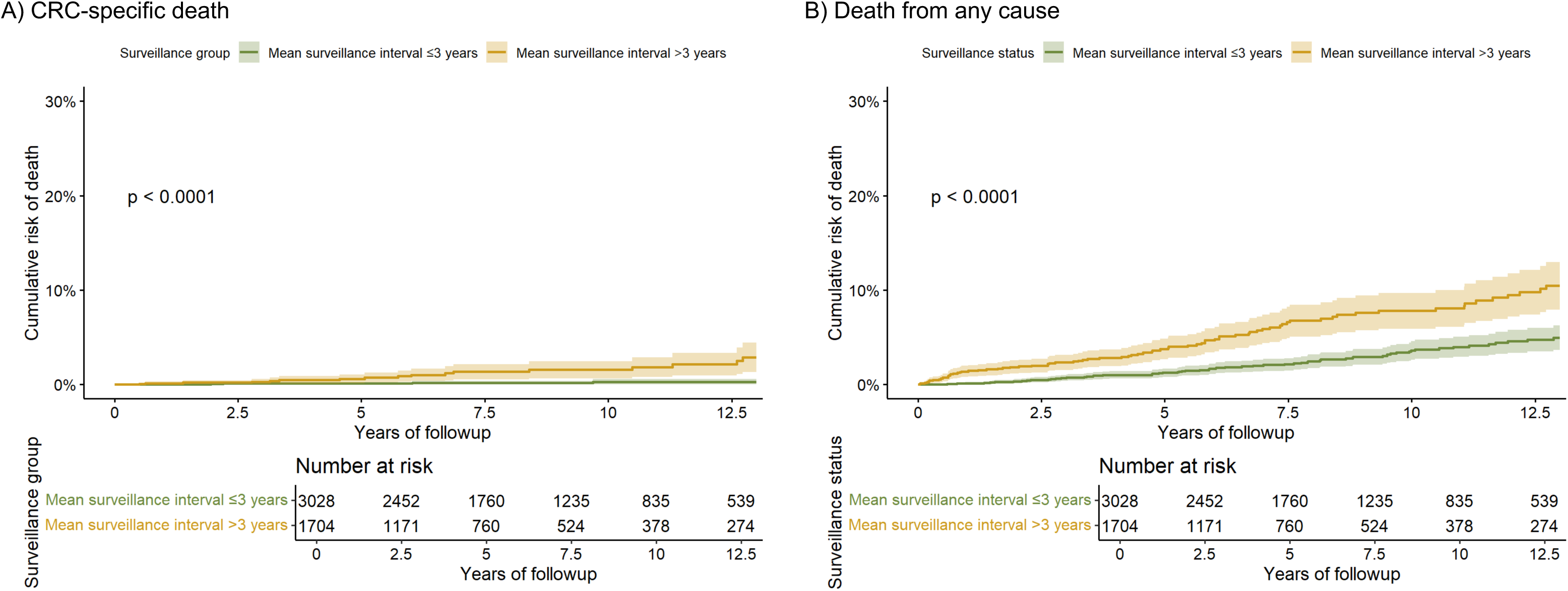
Cumulative risk of death from A) colorectal cancer and B) any cause in MMR pathogenic variant carriers with a mean surveillance interval. ≥ **3 years compared to MMR pathogenic variant carriers with a mean surveillance interval of > 3 years** CRC-specific deaths are defined as any death where a CRC ICD-10 code (C18, C19, C20) appears as a cause code in any section (1A, 1B, 1C, or 2) of the death certificate. A total of 22 CRC-specific deaths occurred in the cohort (5 in the ≤ 3 years interval group, 17 in the > 3 years group). A total of 148 deaths from any cause occurred in the cohort (68 in the ≤ 3 years interval group, 80 in the > 3 years group). P-values are the result of the log rank test.

There was a lower risk of death from any cause amongst individuals who were *‘3-year-adherent’* compared to those who were not (p<0.0001) (**Error! Reference source not found.**). There were 68 deaths amongst 3,028 *‘3-year-adherent’* MMR-carriers (2.2%) and 80 deaths amongst 1,704 non-*’3-year-adherent’* MMR-carriers (4.7%). The cumulative risk of death at 5 and 10 years was 1.25% (95%CI 0.80-1.70%) and 3.61% (95%CI 2.63-4.58%) for those who were *‘3-year-adherent’* and 3.75% (95%CI 2.65-4.84%) and 7.81% (95%CI 5.89-9.69%) for those who were not (Supplementary Table 15). Following adjustment for MMR gene, sex, age at follow up start, and IMD quintile, an MSVI ≤ 3 years was associated with a reduction in risk of death from any cause (HR 0.44, 95%CI 0.31-0.61) (Supplementary Table 16).

There was thus a lower risk of non-CRC death amongst individuals who were *‘3-year-adherent’* compared to those who were not (p<0.0001). There were 63 non-CRC deaths amongst 3,028 *‘3-year-adherent’* MMR-carriers (2.1%) and 63 deaths amongst 1,704 non-*’3-year-adherent’* MMR-carriers (3.7%). The cumulative risk of non-CRC death at 5 and 10 years was 1.14% (95%CI 0.71-1.57%) and 3.33% (95%CI 2.38-4.26%) for those who were *‘3-year-adherent’* and 3.16% (95%CI 2.15-4.16%) and 6.33% (95%CI 4.59-8.04%) for those who were not.

### Sensitivity analyses

To mitigate the impact of short follow up periods on CRC incidence results, we performed sensitivity analysis using the cohort of 4,129 individuals who had at least one year of follow up (603 individuals excluded, Supplementary Table 17). There were 254 CRC diagnoses (Supplementary Table 18). Comparison of AS-AIRs, cumulative incidence rates, and stage distributions produced similar, though attenuated, results, with fewer statistically significant differences (Supplementary Tables 19, 20, and 21).

## DISCUSSION

The uptake and frequency of colonoscopy aligns with previously published regional estimates of adherence amongst MMR-carriers in England and internationally, which range from 50-80%^23–25^. Using a MSVI of ≤ 3 years as a standard of adherence, increased age was associated with a lower likelihood of adherence, while being of white ethnicity or living in a lower deprivation quintile was associated with a higher likelihood, findings which are consistent with previous literature describing participation in population-level national CRC surveillance programmes in England and abroad^26,27^.

It is generally assumed that regular colonoscopies with polypectomies would reduce the incidence of colorectal cancer, as demonstrated in other at-risk populations. However, in these analyses, there was no clear reduction in CRC incidence for MMR-carriers who were *‘3-year-adherent’* compared to ‘*non*-*adherent’* individuals. Failure of surveillance colonoscopy to effectively prevent CRC development may be explained by i) an accelerated adenoma-carcinoma sequence leading to interval cancers, ii) poor quality colonoscopy, or iii) an alternative mechanism(s) of cancer development in LS which is not amenable to secondary prevention via endoscopy.

Even in the absence of CRC prevention (and a consequent reduction in CRC incidence), more frequent colonoscopy potentially could result in diagnosis of CRCs at an earlier stage than they would otherwise have been (altered stage distribution, or ‘stage-shift’). Detection of CRC at earlier stage may yield mortality reduction, whilst also requiring less aggressive clinical management. A change in the relative stage distribution of CRCs, for example an increase in the relative proportion of stage 1 and 2 cancers, may be indicative of stage-shift, but may also be influenced by underlying changes in CRC incidence, and thus does not robustly demonstrate that stage-shift is occurring. Indeed, the true presence of stage-shift can only be robustly demonstrated by evidence of an increase in the rate of diagnosis of early-stage cancers alongside a decrease in the rate of late-stage cancers, indicating that not only have more early-stage cancers been identified, but also that they have been prevented from developing into late-stage cancers.

In ‘*3-year-adherent’* MMR-carriers, there was a change in stage distribution that was statistically significant for *MLH1*-carriers (p=0.024). However, on analysis of stage-specific AS-AIRs no significant changes were observed for any gene.

When tightening the definition of adherence to MSVI ≤ 2 years, in those having more frequent colonoscopies (*‘2-year-adherent’)* there was a *higher* overall cumulative risk of CRC compared to ‘*non*-*adherent’* individuals, with a higher incidence of CRCs observed in most age groups. Comparison of ‘*2-year-adherent’* and ‘*non-2-year-adherent’* MMR-carriers demonstrated both a change in the distribution of CRC stage at diagnosis (p=0.040), with higher AS-AIRs for early-stage CRCs in more frequently surveilled individuals. However, there was no corresponding decrease in late-stage AS-AIRs observed.

This pattern may indicate that an increased number of early-stage cancers have been identified, but that the follow up period was not sufficient for this to be reflected in late-stage cancers. Alternatively, this pattern would be consistent with overdiagnosis, namely that surveillance is detecting cancers which would not otherwise have progressed to a late stage on account of being indolent or transitory. These results are consistent with findings from the PLSD, noting that PLSD examined CRC incidence according to the surveillance recommendations by area of residence, not by individual-level uptake^28^. Notably, we were unable to explore factors which may have a bearing on the risks and benefits of earlier diagnosis, such as rates of cure, organ preservation, or any adverse events or morbidity associated both with treatment and with lack of treatment.

*‘3-year-adherence’* was associated with a significant reduction in the risk of death from CRC and from any cause, even after adjustment for MMR gene, sex, age at start of follow up period, and IMD quintile. The reduction in risk of death from any cause may indicate a possible impact of colonoscopy on CRC-specific survival. However, in the context of a non-randomised study the observed reductions in both CRC-specific and non-CRC-related mortality in the surveilled group may be indicative of fundamental differences between analysis groups, whereby healthier individuals were more likely to participate in surveillance (the ‘healthy screenee effect’). Length and lead bias may also be present. Notably, it might be hypothesised that these same uncaptured socioeconomic and behavioural factors in a group compliant with screening would correlate with behavioural and environmental exposures reducing their risk of CRCs; this adds further weight to the observation of higher CRC incidence in surveillance-compliant groups.

To our knowledge, this is the first large study to directly compare CRC outcomes in MMR-carriers with regular recorded colonoscopic surveillance to those without. The primary limitation of this work is its observational nature: it is thus subject to the limitations of a non-randomised approach. Whilst known relevant covariates were incorporated in the regression analyses, uncaptured confounding differences between those compliant and non-complaint with surveillance cannot be accounted for and are likely to have substantial impact. There is potential for misclassification in the approach used to define analysis groups: the MSVI is a mean and thus does not capture nuance in the distribution of surveillance colonoscopies in each individual, particularly in individuals with short follow up periods (though this is likely mitigated by grouping colonoscopies into 90-day events). Individuals dying or developing a CRC quickly will be classified as non-adherent, which may bias results towards surveillance benefits. To account for these influences, sensitivity analyses were conducted in a cohort of MMR-carriers with ≥ 1 year of follow-up, which produced similar results to the main analyses.

This analysis was limited by our ability to distinguish robustly between surveillance colonoscopies and colonoscopies conducted for other purposes (e.g. as part of a clinical diagnosis or in response to symptoms). While procedures definitely not constituting surveillance events were removed a priori, this is unlikely to have comprehensively removed all non-surveillance colonoscopies, which may impact our findings if symptomatic individuals are more likely to receive additional colonoscopies. Reported colonoscopic findings were not available but may potentially influence clinician surveillance recall timing. Similarly, information on colonoscopy quality and findings was not available, thus we were unable to include factors such as bowel preparation, withdrawal time, or the identification and management of lesions in our analyses. Such factors which influence the effectiveness of surveillance, but were not taken into account, thus may confound results. While colonoscopy quality data is not available for our study period, work to prospectively evaluate the quality of colonoscopy has recently been initiated, but remains at an early stage^29^. Family cancer history was not available but may be an additional covariate influencing adherence to colonoscopy recommendations, and possibly even clinician recall timing. We did not make specific adjustments in our analyses for variations in colonoscopy activity consequent from the Covid-19 pandemic^30^. Private colonoscopies or colonoscopies conducted outside of England may not be captured in HES, and thus are not included.

In this study, we used linked electronic health data to assess the adherence of surveillance colonoscopies in the English LS population to national guidelines, and to determine the impact of regular surveillance on CRC incidence as well as CRC-specific and overall mortality. To our knowledge, this is the first national study of LS surveillance adherence in England, and the largest study internationally to date of CRC outcomes in MMR-carriers analysing records of colonoscopies.

## Contributors

CH and CT designed the analyses. CH accessed the underlying data, coded the analyses, and generated tables and figures for presentation. CM extracted data from the ENLSR. TR performed linkage of LS data to HES. PP, KM, AA, MT, RH, and EM provided critical feedback to inform the research and analysis. BT provided project management for data access and analyses. DT, LP, and SV provided data expertise. CH and CT wrote the manuscript. All authors reviewed the manuscript. CT is the guarantor for this work.

## Data sharing statement

Summary data relevant to the study are included in the article or supplementary information. Individual-level data used in this study are held within NDRS under robust privacy, security and confidentiality procedures and only available to properly authorised analysts and researchers under access arrangements through the NHS England Data Access and Release Service.

## Ethics

Data used in these analyses are collected and stored securely within NDRS under the legal permissions afforded by Section 254 of the Health and Social Care Act 2012, which permits the collection of patient data without requiring informed consent. This analysis was approved by the NDRS analytical project panel (WP00481).

## Acknowledgements

CH is supported by a Wellcome Trust Clinical Research Training Fellowship (Ref 203924/Z/16/Z). CT, LL, EM, SA, BT, MT, ACA and PDP acknowledge grant support from Cancer Research UK (C8620/A8372). KM is supported by a research grant from the charity 40tude Curing Colon Cancer. MT is supported by the NIHR Cambridge Biomedical Research Centre (NIHR203312). ACA is supported by Cancer Research UK (SEBCD3-2024/100001). This work uses data that has been provided by patients and collected by the NHS as part of their care and support. The data are collated, maintained and quality assured by the National Disease Registration Service, which is part of NHS England.

