## Supplementary Material for "Original research: Impact of surveillance colonoscopy on colorectal cancer incidence and mortality in Lynch syndrome - a national observational cohort study of patients in the English NHS 2010-2022"

### Supplementary Materials

3.10. Supplementary table 10: Age-specific five-year annual incidence rates for colorectal cancer for MMR pathogenic variant carriers by mean surveillance interval (≤ 2 years vs > 2 years) 22

3.18. Supplementary table 18: Sensitivity analyses -colorectal cancer diagnoses and stage distribution in a) the whole cohort, and b) analysis groups, for MMR pathogenic variant carriers... 31

### 1. Supplementary methods

#### 1.1. Data sources

##### 1.1.1. National Disease Registration Service datasets

The National Disease Registration Service (NDRS) is the organisation which is responsible for collecting data on cancer, congenital anomalies, and rare diseases in England. NDRS comprises two parallel disease registration services: the National Congenital Anomaly and Rare Disease Registration Service (NCARDRS), responsible for collecting data on congenital anomalies and rare diseases, and the National Cancer Registration and Analysis Service (NCRAS), responsible for collecting data on cancer. In this work, data is sourced from the National Cancer Registration Dataset (NCRD), Germline Genetic Testing dataset (GGT), and Somatic Molecular Testing dataset (SMT) maintained by NCRAS, and the English National Lynch Syndrome Registry (ENLSR) maintained by NCARDRS.

###### 1.1.1.1. Legal and regulatory framework for data collection in the National Disease Registration Service

In England, the NDRS has a special legal instruction from the Secretary of State for Health and Social Care (the National Disease Registries Directions, issued in 2021) to collect specific data on cancer and rare diseases under Section 254 of the Health and Social Care Act 2012[1,2]. Prior to 2021, the NDRS was granted permission to collect data under Section 251 of the National Health Services Act 2006[2].

###### 1.1.1.2. National Cancer Registration Dataset

The National Cancer Registration Dataset (NCRD) is England's population-based cancer registry. It includes information on all primary tumours diagnosed each year in England (International Statistical Classification of Diseases 10$^{th}$ Revision [ICD-10] codes C00-97, D00-48), excluding benign tumours outside the central nervous system[3]. Data collection began in 1971, became nationally complete and inter-operable from 1995, and at time of writing extends through the end of 2022. Data is collected from multiple sources (including patient administration systems, Multidisciplinary Team (MDT) meeting records, oncology records, pathology laboratory reports, diagnostic imaging reports, treatment records, and death certificates), following which it is reviewed, extracted, and re-formatted by trained Cancer

Registration Officers (CROs), who feed the data into a live application (the English National Cancer Online Registration Environment, ENCORE) which processes the data and stores it in an Oracle database[4]. Each individual tumour and each individual patient is given its own unique identifier (Tumour_ID and Patient_ID, respectively), thus the data may be interrogated at the level of the tumour or the patient. The NHS number is the primary patient identifier, but patient identification may also be achieved using date of birth, name, or address[4]. External linkage to the NCRD can be achieved using NHS number or a combination of name, date of birth, and address. For linkage of internal datasets (datasets stored in the NDRS Cancer Analysis System and/or maintained by NCRAS), the Patient_ID or Tumour_ID may be used as linkage keys.

###### 1.1.1.3. Germline Genetic Testing Dataset

The Germline Genetic Testing dataset (GGT) contains information on germline genetic testing carried out within the NHS at regional molecular genetics laboratories since 1999[5]. The main patient identifier is the Pseudo_ID, a unique, one-way, reproducible pseudonym assigned to each individual (no other patient identifiers are held). Due to pseudonymisation of the data, GGT data cannot be linked directly to external datasets, but can be linked to the NCRD using the Pseudo_ID, from which personal identifiers can be obtained allowing linkage to other internal and external datasets.

###### 1.1.1.4. English National Lynch Syndrome Registry

The English National Lynch Syndrome Registry (ENLSR) is a nationally comprehensive registry of MMR PV carriers in England. The ENSLR contains demographic and genetic information on all known MMR PV carriers, from the earliest diagnoses made in clinical genetics services to the present day. Curated and quality assured by NCARDRS, the ENLSR operates at the patient level and can be linked to other NDRS datasets using unique personal identifiers, for example, the NHS Number.

##### 1.1.2. Additional NHS data sources

###### 1.1.2.1. Hospital Episode Statistics

Hospital Episode Statistics (HES) is a national dataset which contains information on all hospital activity in England, including admitted patient care (HES APC) since 1989, outpatients (HES OP) since 2003[6]. The data is collected for reimbursement of hospital activity. Included in the dataset are variables such as personal identifiers (including NHS number and date of birth), patient data (age, sex, and ethnicity), clinical data (including dates of admission, diagnoses, and procedures or operations undertaken), and geographical information (including provider details and area of residence)[7]. Procedures are identified using Office of Population Censuses and Services Classification of Interventions and Procedures version 4 (OPCS-4) codes, each of which correspond to a specific intervention. The personal identifiers used in HES include the NHS Number, which can be used to link to other NHS datasets.

#### 1.2. Methods

##### 1.2.1. Study population

We constructed a cohort for analysis from an extract of the full contents of the ENLSR on the 28th of November 2024, comprising the total known LS population in England on that date. From the ENSLR, we obtained demographic (date of birth, person stated gender [hereafter referred to as sex], NHS number, ethnicity, postcode of residence, vital status, date of death) and genetic (MMR gene, date of clinical LS diagnosis) data. Unique individuals were identified using their NHS number. Where duplicate entries with the same NHS number but inconsistent associated data (e.g. date of Lynch syndrome diagnosis) were found, we harmonised and de-duplicated the data using the processes described in Huntley et al[8].

We linked the de-duplicated ENLSR to the GGT via the NCRD, using NHS Number to link from the ENLSR to the NCRD, and the NCRD Pseudo_ID to link from the NCRD to the GGT. From the GGT we obtained molecular dates of LS diagnosis for individuals who did not have a clinical date of LS diagnosis in the ENSLR. We identified molecular dates of LS diagnosis in the GGT by locating MMR gene tests (*MLH1*, *MSH2*, *MSH6*, *PMS2*, or *EPCAM*) with an abnormal result and a pathogenic variant (pathogenicity class 3, 4, 5, or unknown), and selecting the date associated with the test according to the following hierarchy: date authorised, date requested, date received, and date collected. We imported the date of molecular diagnosis into the ENSLR only where date of clinical diagnosis was missing, and discarded the remaining variables from the GGT.

After cleaning and processing the ENSLR data, we linked unique individuals to the NCRD and HES APC and HES OP datasets using NHS Number. From the NCRD we obtained cancer data (date of diagnosis, type of cancer, stage of cancer at diagnosis), and demographic data (ethnicity, underlying cause of death). From HES we obtained colonoscopy data (type of procedure, date of procedure) and demographic data (ethnicity, IMD quintile [of residence]).

Only procedures matching OPCS-4 codes from a pre-defined list (Supplementary Table 1) were selected from the HES data lake. The codes were identified from a ‘long list’ of OPCS-4 codes used to identify colonoscopies by colleagues in the Post Colonoscopy Colorectal Cancer Audit Project[9], following collaborative review by a consultant gastroenterologist to agree the codes best representing procedures that constituted surveillance events.

Following linkage, we addressed missing ethnicity data in the ENSLR by creating a ‘unified’ ethnicity variable, which combined ethnicity data from three potential data sources: the ENLSR, the NCRD, and HES. For each individual data source, we instated broad ethnicity categories and selected the most recent data submission (i.e. ethnicity linked to a colonoscopy performed in 2020 would be selected over ethnicity linked to a colonoscopy performed in 2015). We then ‘assigned’ each individual a ‘unified ethnicity variable’ according to a hierarchy of the ENLSR, NCRD, and HES (i.e. if the ethnicity category was missing in the ENLSR, it would be taken from NCRD, and so on).

Carriers of PVs in the *EPCAM* gene were included in the *MSH2* category.

##### 1.2.2. Colonoscopy analyses

We combined Individual OPCS-4 codes representing specific colonoscopic procedures into ‘surveillance events’ by grouping all procedures that occurred within 90 days of each other and assigning each surveillance event the date of the first procedure. We calculated the total colonoscopies per person, the mean number of colonoscopies per person, and the mean surveillance interval (total follow up time / number of colonoscopies, MSVI) for each person. We considered an individual’s first colonoscopy adherent to guidelines if it took place within 12 months of the individual becoming eligible for colonoscopic surveillance. We considered a subsequent colonoscopies adherent if it took place within 36 months (3 years) of the previous colonoscopy (3YR_adherent).

With regard to logistic regression, univariable models were constructed for each explanatory variable in turn, and all that were significantly associated with the outcome ($p<0.05$) were carried through to the multivariable model. We calculated p-values using the Wald test and p-values for linear trend in ordinal variables using linear polynomial contrasts.

##### 1.2.3. Colorectal cancer incidence analyses

Confidence intervals for AS-AIRs were estimated using Byar's approximation, and we tested for a statistically significant difference in the AIRs from comparator groups using a two-tailed Z-test.

We estimated cumulative risks of CRC from age 25-74 using the life table method. We used one-year annual incidence rates to estimate the probability of *not* developing cancer in each interval, followed by the cumulative probability of remaining cancer free, and finally converted this to cumulative risk. We assumed zero incidence of CRC before the age of 25 for *MLH1* and *MSH2* PV carriers, and the age of 35 for *MSH6* and *PMS2* PV carriers. We used Greenwood's formula to calculate the standard error for each estimate, and hence the 95% confidence interval.

We tested for a statistically significant difference in the stage distributions of tumours using the chi-square test or Fisher's exact test, depending on the frequencies of events.

##### 1.2.4. Mortality analyses

We used the log-rank test to test for a statistically significant difference in the cumulative risk of death (calculated using Kaplan-Meier methods) between comparator groups.

### 2. Supplementary figures

#### 2.1. Supplementary figure 1

**Figure 1. Cohort assembly**

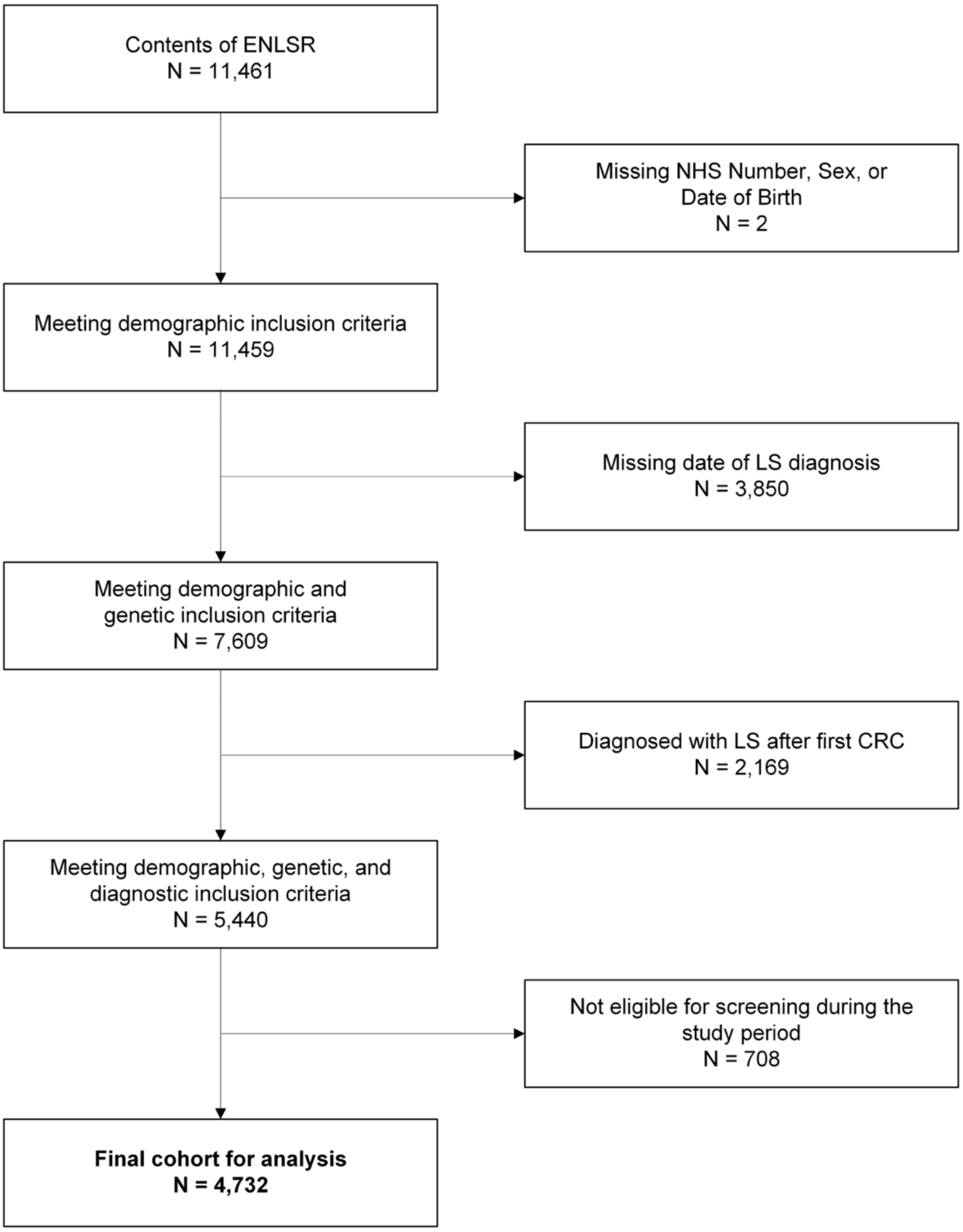

*ENLSR, English National Lynch Syndrome Registry; LS, Lynch syndrome; CRC, colorectal cancer.*

### 3. Supplementary tables

#### 3.1. Supplementary table 1: OPCS-4 codes used to identify surveillance colonoscopies in Hospital Episodes Statistics (HES) data

| OPCS4 | High level descriptor | Low level descriptor |
|---|---|---|
| G791 | Therapeutic endoscopic operations on ileum | Endoscopic extirpation of lesion of ileum |
| G792 | Therapeutic endoscopic operations on ileum | Endoscopic dilation of ileum |
| G793 | Therapeutic endoscopic operations on ileum | Endoscopic insertion of tubal prosthesis into ileum |
| G798 | Therapeutic endoscopic operations on ileum | Other specified |
| G799 | Therapeutic endoscopic operations on ileum | Unspecified |
| G801 | Diagnostic endoscopic examination of ileum | Diagnostic endoscopic examination of ileum and biopsy of lesion of ileum |
| G802 | Diagnostic endoscopic examination of ileum | Wireless capsule endoscopy |
| G808 | Diagnostic endoscopic examination of ileum | Other specified |
| G809 | Diagnostic endoscopic examination of ileum | Unspecified |
| H201 | Endoscopic extirpation of lesion of colon | Fibreoptic endoscopic snare resection of lesion of colon |
| H202 | Endoscopic extirpation of lesion of colon | Fibreoptic endoscopic cauterisation of lesion of colon |
| H203 | Endoscopic extirpation of lesion of colon | Fibreoptic endoscopic laser destruction of lesion of colon |
| H204 | Endoscopic extirpation of lesion of colon | Fibreoptic endoscopic destruction of lesion of colon NEC |
| H205 | Endoscopic extirpation of lesion of colon | Fibreoptic endoscopic submucosal resection of lesion of colon |
| H206 | Endoscopic extirpation of lesion of colon | Fibreoptic endoscopic resection of lesion of colon NEC |
| H207 | Endoscopic extirpation of lesion of colon | Fibreoptic endoscopic mucosal resection of lesion of colon |
| H208 | Endoscopic extirpation of lesion of colon | Other specified |
| H209 | Endoscopic extirpation of lesion of colon | Unspecified |
| H211 | Other therapeutic endoscopic operations on colon | Fibreoptic endoscopic dilation of colon |
| H212 | Other therapeutic endoscopic operations on colon | Fibreoptic endoscopic coagulation of blood vessel of colon |
| H213 | Other therapeutic endoscopic operations on colon | Fibreoptic endoscopic removal of foreign body from colon |
| H214 | Other therapeutic endoscopic operations on colon | Fibreoptic endoscopic insertion of expanding metal stent into colon |
| H215 | Other therapeutic endoscopic operations on colon | Fibreoptic endoscopic decompression of colon |
| H218 | Other therapeutic endoscopic operations on colon | Other specified |
| H219 | Other therapeutic endoscopic operations on colon | Unspecified |
| H221 | Diagnostic endoscopic examination of colon | Diagnostic fibreoptic endoscopic examination of colon and biopsy of lesion of colon |
| H228 | Diagnostic endoscopic examination of colon | Other specified |
| H229 | Diagnostic endoscopic examination of colon | Unspecified |
| H231 | Endoscopic extirpation of lesion of lower bowel using fibreoptic sigmoidoscope | Endoscopic snare resection of lesion of lower bowel using fibreoptic sigmoidoscope |
| H232 | Endoscopic extirpation of lesion of lower bowel using fibreoptic sigmoidoscope | Endoscopic cauterisation of lesion of lower bowel using fibreoptic sigmoidoscope |
| H233 | Endoscopic extirpation of lesion of lower bowel using fibreoptic sigmoidoscope | Endoscopic laser destruction of lesion of lower bowel using fibreoptic sigmoidoscope |
| H234 | Endoscopic extirpation of lesion of lower bowel using fibreoptic sigmoidoscope | Endoscopic destruction of lesion of lower bowel using fibreoptic sigmoidoscope NEC |

| | | |
|---|---|---|
| H235 | Endoscopic extirpation of lesion of lower bowel using fibreoptic sigmoidoscope | Endoscopic submucosal resection of lesion of lower bowel using fibreoptic sigmoidoscope |
| H236 | Endoscopic extirpation of lesion of lower bowel using fibreoptic sigmoidoscope | Endoscopic resection of lesion of lower bowel using fibreoptic sigmoidoscope NEC |
| H238 | Endoscopic extirpation of lesion of lower bowel using fibreoptic sigmoidoscope | Other specified |
| H239 | Endoscopic extirpation of lesion of lower bowel using fibreoptic sigmoidoscope | Unspecified |
| H241 | Other therapeutic endoscopic operations on lower bowel using fibreoptic sigmoidoscope | Endoscopic dilation of lower bowel using fibreoptic sigmoidoscope |
| H242 | Other therapeutic endoscopic operations on lower bowel using fibreoptic sigmoidoscope | Endoscopic coagulation of blood vessel of lower bowel using fibreoptic sigmoidoscope |
| H243 | Other therapeutic endoscopic operations on lower bowel using fibreoptic sigmoidoscope | Endoscopic insertion of tubal prosthesis into lower bowel using fibreoptic sigmoidoscope |
| H244 | Other therapeutic endoscopic operations on lower bowel using fibreoptic sigmoidoscope | Endoscopic insertion of expanding metal stent into lower bowel using fibreoptic sigmoidoscope |
| H248 | Other therapeutic endoscopic operations on lower bowel using fibreoptic sigmoidoscope | Other specified |
| H249 | Other therapeutic endoscopic operations on lower bowel using fibreoptic sigmoidoscope | Unspecified |
| H251 | Diagnostic endoscopic examination of lower bowel using fibreoptic sigmoidoscope | Diagnostic endoscopic examination of lower bowel and biopsy of lesion of lower bowel using fibreoptic sigmoidoscope |
| H252 | Diagnostic endoscopic examination of lower bowel using fibreoptic sigmoidoscope | Diagnostic endoscopic examination of lower bowel and sampling for bacterial overgrowth using fibreoptic sigmoidoscope |
| H258 | Diagnostic endoscopic examination of lower bowel using fibreoptic sigmoidoscope | Other specified |
| H259 | Diagnostic endoscopic examination of lower bowel using fibreoptic sigmoidoscope | Unspecified |
| H261 | Endoscopic extirpation of lesion of sigmoid colon using rigid sigmoidoscope | Endoscopic snare resection of lesion of sigmoid colon using rigid sigmoidoscope |
| H262 | Endoscopic extirpation of lesion of sigmoid colon using rigid sigmoidoscope | Endoscopic cauterisation of lesion of sigmoid colon using rigid sigmoidoscope |
| H263 | Endoscopic extirpation of lesion of sigmoid colon using rigid sigmoidoscope | Endoscopic laser destruction of lesion of sigmoid colon using rigid sigmoidoscope |
| H264 | Endoscopic extirpation of lesion of sigmoid colon using rigid sigmoidoscope | Endoscopic cryotherapy to lesion of sigmoid colon using rigid sigmoidoscope |
| H265 | Endoscopic extirpation of lesion of sigmoid colon using rigid sigmoidoscope | Endoscopic destruction of lesion of sigmoid colon using rigid sigmoidoscope NEC |
| H266 | Endoscopic extirpation of lesion of sigmoid colon using rigid sigmoidoscope | Endoscopic submucosal resection of lesion of sigmoid colon using rigid sigmoidoscope |
| H267 | Endoscopic extirpation of lesion of sigmoid colon using rigid sigmoidoscope | Endoscopic resection of lesion of sigmoid colon using rigid sigmoidoscope NEC |
| H268 | Endoscopic extirpation of lesion of sigmoid colon using rigid sigmoidoscope | Other specified |
| H269 | Endoscopic extirpation of lesion of sigmoid colon using rigid sigmoidoscope | Unspecified |
| H271 | Other therapeutic endoscopic operations on sigmoid colon using rigid sigmoidoscope | Endoscopic dilation of sigmoid colon using rigid sigmoidoscope |
| H272 | Other therapeutic endoscopic operations on sigmoid colon using rigid sigmoidoscope | Endoscopic removal of foreign body from sigmoid colon using rigid sigmoidoscope |
| H273 | Other therapeutic endoscopic operations on sigmoid colon using rigid sigmoidoscope | Endoscopic insertion of tubal prosthesis into sigmoid colon using rigid sigmoidoscope |
| H274 | Other therapeutic endoscopic operations on sigmoid colon using rigid sigmoidoscope | Endoscopic insertion of expanding metal stent into sigmoid colon using rigid sigmoidoscope |

| | | |
|---|---|---|
| H278 | Other therapeutic endoscopic operations on sigmoid colon using rigid sigmoidoscope | Other specified |
| H279 | Other therapeutic endoscopic operations on sigmoid colon using rigid sigmoidoscope | Unspecified |
| H281 | Diagnostic endoscopic examination of sigmoid colon using rigid sigmoidoscope | Diagnostic endoscopic examination of sigmoid colon and biopsy of lesion of sigmoid colon using rigid sigmoidoscope |
| H288 | Diagnostic endoscopic examination of sigmoid colon using rigid sigmoidoscope | Other specified |
| H289 | Diagnostic endoscopic examination of sigmoid colon using rigid sigmoidoscope | Unspecified |

*Only procedures matching OPCS-4 codes from a pre-defined list were selected from the HES data lake (both outpatient and inpatient). The codes were identified from a 'long list' of OPCS-4 codes used to identify colonoscopies in the Post Colonoscopy Colorectal Cancer Audit Project, following collaborative review by a consultant gastroenterologist to agree the codes best representing procedures that constituted surveillance events. Individual OPCS-4 codes representing specific colonoscopic procedures were combined into 'surveillance events' by grouping all procedures that occurred within 90 days of each other and assigning each surveillance event the date of the first procedure. OPCS-4, Office of Population Censuses and Surveys Classification of Interventions and Procedures version 4; HES, Hospital Episodes Statistics.*

#### 3.2. Supplementary table 2: Years of follow up for colonoscopy, incidence, and mortality analyses

| | Colonoscopy / incidence analyses | | Mortality analysis | |
|---|---|---|---|---|
| | **Total** | **Median** | **Total** | **Median** |
| **MLH1** | 8610 | 5.75 | 9363 | 6.62 |
| **MSH2** | 12280 | 5.95 | 13328 | 6.76 |
| **MSH6** | 4875 | 4.03 | 5191 | 4.17 |
| **PMS2** | 2102 | 3.33 | 2208 | 3.51 |
| **All genes** | 27866 | 4.88 | 30091 | 5.55 |

*Colonoscopy and incidence analyses have the same follow up period. 'All genes' refers to all MMR genes combined.*

#### 3.3. Supplementary table 3: Distribution of surveillance colonoscopies in the Lynch syndrome cohort and adherence to national colonoscopic surveillance guidelines

| | | **All genes** | | **MLH1** | | **MSH2** | | **MSH6** | | **PMS2** | |
|---|---|---|---|---|---|---|---|---|---|---|---|
| | | **n** | **%** | **n** | **%** | **n** | **%** | **n** | **%** | **n** | **%** |
| **Totals** | Total individuals | 4732 | | 1315 | | 1856 | | 1011 | | 550 | |
| | Total colonoscopies | 11201 | | 3412 | | 4972 | | 2015 | | 802 | |
| | Mean colonoscopies per person | 2.37 | | 2.59 | | 2.68 | | 1.99 | | 1.46 | |
| | Mean surveillance interval | 2.51 | | 2.58 | | 2.61 | | 2.36 | | 2.22 | |
| **First colonoscopies** | Total first colonoscopies | 3788 | | 1074 | | 1541 | | 794 | | 379 | |
| | First colonoscopy adherent | 2492 | 65.8 | 646 | 60.1 | 984 | 63.9 | 574 | 72.3 | 288 | 76.0 |
| | First colonoscopy not adherent | 1296 | 34.2 | 428 | 39.9 | 557 | 36.1 | 220 | 27.7 | 91 | 24.0 |
| | Median time to first colonoscopy (days) | 214.5 | | 265 | | 230 | | 184 | | 167 | |
| **Subsequent colonoscopies** | Total subsequent colonoscopies | 7413 | | 2338 | | 3431 | | 1221 | | 423 | |
| | Subsequent colonoscopy within 24 months | 3053 | 41.2 | 977 | 41.8 | 1417 | 41.3 | 477 | 39.1 | 182 | 43.0 |
| | Subsequent colonoscopy within 36 months | 6760 | 91.2 | 2122 | 90.8 | 3158 | 92.0 | 1097 | 89.8 | 383 | 90.5 |

*'Surveillance colonoscopies and compliance with surveillance guidelines in the full cohort (no limits on follow up time). Surveillance colonoscopies were identified from a list of OPCS-4 codes, combined into a single event if occuring within 90 days of each other, and assigned the date of the first code in the event.*

#### 3.4. Supplementary table 4: Number of first colorectal cancer diagnoses and stage at diagnosis broken down by MMR gene

| | | | n | % |
|---|---|---|---|---|
| **MLH1** | CRC outcome | CRC diagnosis | 138 | 10.5 |
| | | No CRC diagnosis | 1177 | 89.5 |
| | Stage at diagnosis | Stage 1 | 54 | 39.4 |
| | | Stage 2 | 39 | 28.5 |
| | | Stage 3 | 27 | 19.7 |
| | | Stage 4 | 5 | 3.6 |
| | | Unknown | 12 | 8.8 |
| **MSH2** | CRC outcome | CRC diagnosis | 167 | 9.0 |
| | | No CRC diagnosis | 1689 | 91.0 |
| | Stage at diagnosis | Stage 1 | 70 | 41.9 |
| | | Stage 2 | 37 | 22.2 |
| | | Stage 3 | 38 | 22.8 |
| | | Stage 4 | 4 | 2.4 |
| | | Unknown | 18 | 10.8 |
| **MSH6** | CRC outcome | CRC diagnosis | 23 | 2.3 |
| | | No CRC diagnosis | 988 | 97.7 |
| | Stage at diagnosis | Stage 1 | 8 | 34.8 |
| | | Stage 2 | 7 | 30.4 |
| | | Stage 3 | 4 | 17.4 |
| | | Stage 4 | 1 | 4.3 |
| | | Unknown | 3 | 13.0 |
| **PMS2** | CRC outcome | CRC diagnosis | 6 | 1.1 |
| | | No CRC diagnosis | 544 | 98.9 |
| | Stage at diagnosis | Stage 1 | 3 | 50.0 |
| | | Stage 2 | 0 | 0.0 |
| | | Stage 3 | 1 | 16.7 |
| | | Stage 4 | 1 | 16.7 |
| | | Unknown | 1 | 16.7 |
| **All genes** | CRC outcome | CRC diagnosis | 334 | 7.1 |
| | | No CRC diagnosis | 4398 | 92.9 |
| | Stage at diagnosis | Stage 1 | 135 | 40.4 |
| | | Stage 2 | 83 | 24.9 |
| | | Stage 3 | 70 | 21.0 |
| | | Stage 4 | 11 | 3.3 |
| | | Unknown | 35 | 10.5 |

*Denominators for the percentages in colorectal cancer outcome rows are the group totals. Denominators for the percentages in the stage at diagnosis rows are the numbers of colorectal cancers in that specific group. p-values are calculated using the Chi-square test or Fisher's Exact Test. Tumours are staged at diagnosis. CRC, colorectal cancer; all genes, all MMR genes combined.*

#### 3.5. Supplementary table 5: Number of first colorectal cancer diagnoses and stage at diagnosis for MLH1, MSH2, MSH6, and PMS2 PV carriers, by mean surveillance interval (less than or equal to 3 years vs greater than 3 years)

| | | | Mean surveillance interval | | | | |
|---|---|---|---|---|---|---|---|
| | | | ≤ 3 years | | > 3 years | | |
| | | | n | % | n | % | p-value |
| MLH1 | CRC outcome | CRC diagnosis | 97 | 11.6 | 41 | 8.6 | |
| | | No CRC diagnosis | 740 | 88.4 | 437 | 91.4 | |
| | Stage at diagnosis | Stage 1 | 43 | 44.3 | 11 | 26.8 | 0.024 |
| | | Stage 2 | 21 | 21.6 | 18 | 43.9 | |
| | | Stage 3 | 18 | 18.6 | 9 | 22.0 | |
| | | Stage 4 | 3 | 3.1 | 2 | 4.9 | |
| | | Unknown | 12 | 12.4 | 1 | 2.4 | |
| MSH2 | CRC outcome | CRC diagnosis | 118 | 9.6 | 49 | 7.8 | |
| | | No CRC diagnosis | 1106 | 90.4 | 583 | 92.2 | |
| | Stage at diagnosis | Stage 1 | 54 | 45.8 | 16 | 32.7 | 0.491 |
| | | Stage 2 | 25 | 21.2 | 12 | 24.5 | |
| | | Stage 3 | 25 | 21.2 | 13 | 26.5 | |
| | | Stage 4 | 2 | 1.7 | 2 | 4.1 | |
| | | Unknown | 12 | 10.2 | 6 | 12.2 | |
| MSH6 | CRC outcome | CRC diagnosis | 17 | 2.6 | 6 | 1.7 | |
| | | No CRC diagnosis | 638 | 97.4 | 350 | 98.3 | |
| | Stage at diagnosis | Stage 1 | 7 | 41.2 | 1 | 16.7 | 0.693 |
| | | Stage 2 | 4 | 23.5 | 3 | 50.0 | |
| | | Stage 3 | 3 | 17.6 | 1 | 16.7 | |
| | | Stage 4 | 1 | 5.9 | 0 | 0.0 | |
| | | Unknown | 2 | 11.8 | 1 | 16.7 | |
| PMS2 | CRC outcome | CRC diagnosis | 1 | 0.3 | 5 | 2.1 | |
| | | No CRC diagnosis | 311 | 99.7 | 233 | 97.9 | |
| | Stage at diagnosis | Stage 1 | 0 | 0.0 | 3 | 60.0 | 0.500 |
| | | Stage 2 | 0 | 0.0 | 0 | 0.0 | |
| | | Stage 3 | 1 | 100.0 | 0 | 0.0 | |
| | | Stage 4 | 0 | 0.0 | 1 | 20.0 | |
| | | Unknown | 0 | 0.0 | 1 | 20.0 | |
| All genes | CRC outcome | CRC diagnosis | 233 | 7.7 | 101 | 5.9 | |
| | | No CRC diagnosis | 2795 | 92.3 | 1603 | 94.1 | |
| | Stage at diagnosis | Stage 1 | 104 | 44.6 | 31 | 30.7 | 0.068 |
| | | Stage 2 | 50 | 21.5 | 33 | 32.7 | |
| | | Stage 3 | 47 | 20.2 | 23 | 22.8 | |
| | | Stage 4 | 6 | 2.6 | 5 | 5.0 | |
| | | Unknown | 26 | 11.2 | 9 | 8.9 | |

*Denominators for the percentages in colorectal cancer outcome rows are the group totals. Denominators for the percentages in the stage at diagnosis rows are the numbers of colorectal cancers in that specific group. p-values are calculated using the Chi-square test or Fisher's Exact Test. Tumours are staged at diagnosis. CRC, colorectal cancer; all genes, all MMR genes combined.*

#### 3.6. Supplementary table 6: Age-specific five-year annual incidence rates for colorectal cancers for MMR pathogenic variant carriers by mean surveillance interval (≤ 3 years vs > 3 years)

| | Age | Mean surveillance interval: 3 years | | | | | | | | | | |
|---|---|---|---|---|---|---|---|---|---|---|---|---|
| | | ≤ 3 years | | | | | > 3 years | | | | | |
| | | No. of CRCs | Obs Years | AS-AIR | Lower CI | Upper CI | No. of CRCs | Obs Years | AS-AIR | Lower CI | Upper CI | p-value |
| MLH1 | 25-29 | 4 | 702 | 569 | 155 | 1458 | 4 | 382 | 1048 | 285 | 2683 | 0.492 |
| | 30-34 | 7 | 886 | 790 | 317 | 1627 | 2 | 358 | 558 | 68 | 2016 | 0.699 |
| | 35-39 | 15 | 822 | 1826 | 1021 | 3011 | 2 | 325 | 616 | 75 | 2225 | 0.106 |
| | 40-44 | 24 | 1407 | 1706 | 1093 | 2538 | 5 | 532 | 939 | 305 | 2192 | 0.206 |
| | 50-54 | 10 | 648 | 1543 | 739 | 2838 | 6 | 301 | 1996 | 733 | 4345 | 0.670 |
| | 55-59 | 14 | 526 | 2664 | 1455 | 4469 | 2 | 303 | 661 | 80 | 2386 | 0.039 |
| | 60-64 | 13 | 400 | 3251 | 1729 | 5559 | 9 | 283 | 3180 | 1454 | 6037 | 0.963 |
| | 65-69 | 6 | 265 | 2266 | 832 | 4932 | 7 | 183 | 3830 | 1540 | 7891 | 0.418 |
| | 70-74 | 4 | 167 | 2397 | 653 | 6138 | 4 | 120 | 3323 | 905 | 8509 | 0.699 |
| MSH2 | 25-29 | 7 | 864 | 810 | 326 | 1669 | 0 | 386 | 0 | 0 | 956 | 0.054 |
| | 30-34 | 11 | 1123 | 980 | 488 | 1753 | 3 | 425 | 706 | 146 | 2062 | 0.640 |
| | 35-39 | 9 | 1089 | 827 | 378 | 1569 | 3 | 387 | 776 | 160 | 2268 | 0.935 |
| | 40-44 | 17 | 2239 | 759 | 442 | 1216 | 12 | 837 | 1435 | 740 | 2506 | 0.170 |
| | 50-54 | 20 | 1041 | 1922 | 1174 | 2969 | 8 | 439 | 1822 | 787 | 3590 | 0.906 |
| | 55-59 | 19 | 871 | 2181 | 1313 | 3407 | 10 | 410 | 2440 | 1168 | 4487 | 0.796 |
| | 60-64 | 18 | 599 | 3005 | 1780 | 4749 | 8 | 329 | 2432 | 1050 | 4792 | 0.638 |
| | 65-69 | 8 | 499 | 1602 | 692 | 3157 | 1 | 267 | 375 | 9 | 2090 | 0.136 |
| | 70-74 | 9 | 274 | 3282 | 1501 | 6231 | 4 | 202 | 1981 | 540 | 5072 | 0.436 |
| MSH6 | 25-29 | | | | | | | | | | | |
| | 30-34 | | | | | | | | | | | |
| | 35-39 | 0 | 312 | 0 | 0 | 1184 | 0 | 178 | 0 | 0 | 2078 | 1.000 |
| | 40-44 | 1 | 848 | 118 | 3 | 657 | 0 | 325 | 0 | 0 | 1136 | 0.724 |
| | 50-54 | 5 | 534 | 937 | 304 | 2187 | 0 | 178 | 0 | 0 | 2069 | 0.189 |
| | 55-59 | 4 | 603 | 664 | 181 | 1699 | 1 | 240 | 416 | 11 | 2317 | 0.725 |
| | 60-64 | 3 | 485 | 618 | 128 | 1807 | 0 | 202 | 0 | 0 | 1823 | 0.328 |
| | 65-69 | 2 | 345 | 580 | 70 | 2094 | 4 | 203 | 1974 | 538 | 5053 | 0.269 |
| | 70-74 | 2 | 265 | 754 | 91 | 2725 | 1 | 157 | 635 | 16 | 3540 | 0.916 |
| PMS2 | 25-29 | | | | | | | | | | | |
| | 30-34 | | | | | | | | | | | |
| | 35-39 | 0 | 144 | 0 | 0 | 2566 | 0 | 117 | 0 | 0 | 3153 | 1.000 |
| | 40-44 | 0 | 319 | 0 | 0 | 1158 | 1 | 183 | 547 | 14 | 3045 | 0.509 |
| | 50-54 | 0 | 210 | 0 | 0 | 1753 | 0 | 84 | 0 | 0 | 4371 | 1.000 |
| | 55-59 | 1 | 212 | 473 | 12 | 2634 | 2 | 109 | 1835 | 222 | 6629 | 0.440 |
| | 60-64 | 0 | 198 | 0 | 0 | 1866 | 0 | 118 | 0 | 0 | 3120 | 1.000 |
| | 65-69 | 0 | 153 | 0 | 0 | 2404 | 2 | 82 | 2439 | 295 | 8811 | 0.280 |
| | 70-74 | 0 | 94 | 0 | 0 | 3915 | 0 | 78 | 0 | 0 | 4711 | 1.000 |
| All genes | 25-29 | 11 | 1566 | 702 | 350 | 1257 | 4 | 768 | 521 | 142 | 1334 | 0.635 |
| | 30-34 | 18 | 2009 | 896 | 531 | 1416 | 5 | 783 | 638 | 207 | 1489 | 0.517 |
| | 35-39 | 24 | 2366 | 1015 | 650 | 1510 | 5 | 1006 | 497 | 161 | 1160 | 0.124 |
| | 40-44 | 42 | 4812 | 873 | 629 | 1180 | 18 | 1877 | 959 | 568 | 1516 | 0.757 |
| | 50-54 | 35 | 2433 | 1439 | 1002 | 2001 | 14 | 1002 | 1397 | 763 | 2344 | 0.930 |
| | 55-59 | 38 | 2211 | 1719 | 1216 | 2359 | 15 | 1062 | 1412 | 790 | 2330 | 0.531 |
| | 60-64 | 34 | 1682 | 2022 | 1400 | 2825 | 17 | 933 | 1823 | 1061 | 2919 | 0.739 |
| | 65-69 | 16 | 1263 | 1267 | 724 | 2058 | 14 | 734 | 1907 | 1042 | 3200 | 0.323 |
| | 70-74 | 15 | 800 | 1874 | 1048 | 3091 | 9 | 558 | 1613 | 738 | 3062 | 0.741 |

*Only first colorectal cancers are included. Follow up for MSH6 and PMS2 begins at age 35, in line with eligibility for colonoscopic surveillance. 95% confidence intervals are calculated using Byar's method. p-values are the result of a two-sided Z-test. AS-AIR, age-specific annual incidence rate per 100,000; CRC, colorectal cancer.*

#### 3.7. Cumulative risk (in %) of colorectal cancer in MMR pathogenic variant carriers, by mean surveillance interval (≤ 3 years vs > 3 years)

| | | **Mean surveillance interval: 3 years** | | | | | |
|---|---|---|---|---|---|---|---|
| | | **≤ 3 years** | | | **> 3 years** | | |
| | **Age** | **Cum risk** | **Lower CI (95%)** | **Upper CI (95%)** | **Cum risk** | **Lower CI (95%)** | **Upper CI (95%)** |
| **MLH1** | 25 | 1.3% | 0.0% | 3.7% | 0.0% | 0.0% | 0.0% |
| | 30 | 6.4% | 0.9% | 11.9% | 3.4% | 0.4% | 6.3% |
| | 35 | 7.7% | 1.6% | 13.8% | 7.1% | 3.1% | 11.1% |
| | 40 | 11.9% | 4.3% | 19.6% | 14.9% | 9.2% | 20.6% |
| | 45 | 13.7% | 5.3% | 22.2% | 21.3% | 14.6% | 28.1% |
| | 50 | 21.2% | 10.6% | 31.8% | 29.1% | 21.2% | 36.9% |
| | 55 | 26.5% | 14.8% | 38.1% | 36.3% | 27.5% | 45.0% |
| | 60 | 31.1% | 18.6% | 43.6% | 43.5% | 33.9% | 53.1% |
| | 65 | 40.6% | 26.6% | 54.6% | 52.9% | 42.1% | 63.7% |
| | 70 | 53.2% | 37.0% | 69.4% | 56.7% | 45.3% | 68.0% |
| | 74 | 57.5% | 40.3% | 74.8% | 62.0% | 49.5% | 74.4% |
| **MSH2** | 25 | 0.0% | 0.0% | 0.0% | 0.0% | 0.0% | 0.0% |
| | 30 | 0.0% | 0.0% | 0.0% | 4.1% | 1.3% | 6.9% |
| | 35 | 3.5% | 0.0% | 7.5% | 9.8% | 5.7% | 13.9% |
| | 40 | 10.6% | 3.8% | 17.4% | 12.8% | 8.1% | 17.4% |
| | 45 | 17.0% | 8.6% | 25.4% | 16.2% | 11.1% | 21.3% |
| | 50 | 21.7% | 12.4% | 31.0% | 20.2% | 14.6% | 25.9% |
| | 55 | 28.6% | 18.2% | 39.0% | 26.3% | 19.9% | 32.6% |
| | 60 | 36.3% | 24.8% | 47.8% | 35.3% | 27.9% | 42.7% |
| | 65 | 43.1% | 30.7% | 55.6% | 44.2% | 35.8% | 52.7% |
| | 70 | 45.5% | 32.6% | 58.4% | 49.7% | 40.6% | 58.9% |
| | 74 | 49.4% | 35.8% | 63.0% | 55.1% | 45.2% | 65.1% |
| **MSH6** | 35 | 0.0% | 0.0% | 0.0% | 0.0% | 0.0% | 0.0% |
| | 40 | 0.0% | 0.0% | 0.0% | 0.0% | 0.0% | 0.0% |
| | 45 | 0.0% | 0.0% | 0.0% | 0.0% | 0.0% | 0.0% |
| | 50 | 0.0% | 0.0% | 0.0% | 2.2% | 0.0% | 5.2% |
| | 55 | 0.0% | 0.0% | 0.0% | 7.2% | 2.2% | 12.2% |
| | 60 | 2.0% | 0.0% | 5.8% | 10.4% | 4.5% | 16.2% |
| | 65 | 6.9% | 0.0% | 14.4% | 11.4% | 5.2% | 17.6% |
| | 70 | 14.1% | 3.2% | 25.0% | 14.3% | 7.0% | 21.7% |
| | 74 | 14.1% | 3.2% | 25.0% | 17.6% | 9.0% | 26.1% |
| **PMS2** | 35 | 0.0% | 0.0% | 0.0% | 0.0% | 0.0% | 0.0% |
| | 40 | 0.0% | 0.0% | 0.0% | 0.0% | 0.0% | 0.0% |
| | 45 | 0.0% | 0.0% | 0.0% | 4.6% | 0.0% | 13.3% |
| | 50 | 0.0% | 0.0% | 0.0% | 4.6% | 0.0% | 13.3% |
| | 55 | 2.3% | 0.0% | 6.8% | 4.6% | 0.0% | 13.3% |
| | 60 | 2.3% | 0.0% | 6.8% | 12.1% | 0.0% | 25.0% |
| | 65 | 2.3% | 0.0% | 6.8% | 12.1% | 0.0% | 25.0% |
| | 70 | 2.3% | 0.0% | 6.8% | 27.3% | 5.6% | 49.0% |
| | 74 | 2.3% | 0.0% | 6.8% | 27.3% | 5.6% | 49.0% |
| **All genes** | 25 | 0.7% | 0.0% | 2.0% | 0.0% | 0.0% | 0.0% |
| | 30 | 3.2% | 0.4% | 6.0% | 3.8% | 1.7% | 5.8% |
| | 35 | 5.7% | 2.0% | 9.3% | 8.5% | 5.6% | 11.3% |
| | 40 | 9.9% | 5.3% | 14.5% | 12.5% | 9.2% | 15.8% |
| | 45 | 13.8% | 8.5% | 19.1% | 16.1% | 12.4% | 19.7% |
| | 50 | 18.2% | 12.3% | 24.1% | 20.6% | 16.6% | 24.7% |
| | 55 | 23.0% | 16.5% | 29.4% | 26.4% | 21.9% | 30.8% |
| | 60 | 28.6% | 21.5% | 35.6% | 32.6% | 27.7% | 37.4% |
| | 65 | 35.1% | 27.5% | 42.8% | 38.9% | 33.6% | 44.2% |
| | 70 | 41.9% | 33.6% | 50.3% | 42.7% | 37.1% | 48.4% |
| | 74 | 44.5% | 35.9% | 53.2% | 47.0% | 40.8% | 53.1% |

*Risk is assumed to be zero below the age of 25 for MLH1 and MSH2 carriers, and below the age of 35 for MSH6 and PMS2 carriers. Cumulative risks are calculated using life table methods, using age-specific annual incidence rates derived from the cohort/analysis groups. Standard errors were calculated using Greenwood's formula, and hence the 95% confidence interval. Cum risk, Cumulative risk; CI, Confidence interval (95%)*

#### 3.8. Supplementary table 8: Stage- and age- specific five-year annual incidence rates for all MMR pathogenic variant carriers by mean surveillance interval (≤ 3 years vs > 3 years)

| | | ≤ 3 years | | | | | > 3 years | | | | | |
|---|---|---|---|---|---|---|---|---|---|---|---|---|
| | | No. of CRCs | Obs Years | AS-AIR | Lower CI (95%) | Upper CI (95%) | No. of CRCs | Obs Years | AS-AIR | Lower CI (95%) | Upper CI (95%) | p-value |
| Stage 1 and 2 | 25-29 | 6 | 1566 | 383 | 141 | 834 | 2 | 768 | 261 | 32 | 941 | 0.675 |
| | 30-34 | 10 | 2009 | 498 | 238 | 915 | 3 | 783 | 383 | 79 | 1119 | 0.717 |
| | 35-39 | 15 | 2366 | 634 | 355 | 1046 | 2 | 1006 | 199 | 24 | 718 | 0.082 |
| | 40-44 | 28 | 4812 | 582 | 387 | 841 | 10 | 1877 | 533 | 255 | 980 | 0.823 |
| | 50-54 | 23 | 2433 | 945 | 599 | 1419 | 9 | 1002 | 898 | 411 | 1705 | 0.903 |
| | 55-59 | 28 | 2211 | 1267 | 841 | 1831 | 9 | 1062 | 847 | 387 | 1609 | 0.296 |
| | 60-64 | 22 | 1682 | 1308 | 820 | 1981 | 11 | 933 | 1180 | 588 | 2111 | 0.792 |
| | 65-69 | 12 | 1263 | 950 | 491 | 1660 | 12 | 734 | 1635 | 844 | 2856 | 0.249 |
| | 70-74 | 10 | 800 | 1249 | 598 | 2298 | 6 | 558 | 1075 | 395 | 2341 | 0.792 |
| Stage 3 and 4 | 25-29 | 4 | 1566 | 255 | 70 | 654 | 2 | 768 | 261 | 32 | 941 | 0.985 |
| | 30-34 | 7 | 2009 | 348 | 140 | 718 | 2 | 783 | 255 | 31 | 922 | 0.731 |
| | 35-39 | 6 | 2366 | 254 | 93 | 552 | 3 | 1006 | 298 | 62 | 872 | 0.851 |
| | 40-44 | 6 | 4812 | 125 | 46 | 271 | 7 | 1877 | 373 | 150 | 769 | 0.139 |
| | 50-54 | 9 | 2433 | 370 | 169 | 702 | 5 | 1002 | 499 | 162 | 1164 | 0.656 |
| | 55-59 | 7 | 2211 | 317 | 127 | 652 | 1 | 1062 | 94 | 2 | 525 | 0.239 |
| | 60-64 | 9 | 1682 | 535 | 245 | 1016 | 6 | 933 | 643 | 236 | 1400 | 0.761 |
| | 65-69 | 2 | 1263 | 158 | 19 | 572 | 0 | 734 | 0 | 0 | 503 | 0.406 |
| | 70-74 | 3 | 800 | 375 | 77 | 1095 | 2 | 558 | 358 | 43 | 1295 | 0.968 |

*Only first cancers are included. Tumours are staged at diagnosis. P-values are the result of a two-tailed Z-test. 95% confidence intervals are calculated using Byar's method. AS-AIR, age-specific annual incidence rate per 100,000 ; CRC, colorectal cancer; LCI, Lower Confidence Interval (95%); MSVI, mean surveillance interval; UCI, Upper Confidence Interval (95%), Years, years of observation.*

#### 3.9. Supplementary table 9: Number of first colorectal cancer diagnoses and stage at diagnosis for MMR pathogenic variant carriers, by mean surveillance interval (≤ 2 years vs > 2 years)

| | | | MSVI ≤ 2 years | | MSVI> 2 years | | |
|---|---|---|---|---|---|---|---|
| | | | n | % | n | % | p-value |
| **MLH1** | CRC outcome | CRC diagnosis | 62 | 15.1 | 76 | 8.4 | |
| | | No CRC diagnosis | 348 | 84.9 | 829 | 91.6 | |
| | Stage at diagnosis | Stage 1 | 29 | 46.8 | 25 | 32.9 | 0.047 |
| | | Stage 2 | 10 | 16.1 | 29 | 38.2 | |
| | | Stage 3 | 13 | 21.0 | 14 | 18.4 | |
| | | Stage 4 | 2 | 3.2 | 3 | 3.9 | |
| | | Unknown | 8 | 12.9 | 5 | 6.6 | |
| **MSH2** | CRC outcome | CRC diagnosis | 81 | 13.3 | 86 | 6.9 | |
| | | No CRC diagnosis | 527 | 86.7 | 1162 | 93.1 | |
| | Stage at diagnosis | Stage 1 | 38 | 46.9 | 32 | 37.2 | 0.666 |
| | | Stage 2 | 16 | 19.8 | 21 | 24.4 | |
| | | Stage 3 | 17 | 21.0 | 21 | 24.4 | |
| | | Stage 4 | 1 | 1.2 | 3 | 3.5 | |
| | | Unknown | 9 | 11.1 | 9 | 10.5 | |
| **MSH6** | CRC outcome | CRC diagnosis | 13 | 3.5 | 10 | 1.6 | |
| | | No CRC diagnosis | 358 | 96.5 | 630 | 98.4 | |
| | Stage at diagnosis | Stage 1 | 6 | 46.2 | 2 | 20.0 | 0.606 |
| | | Stage 2 | 3 | 23.1 | 4 | 40.0 | |
| | | Stage 3 | 2 | 15.4 | 2 | 20.0 | |
| | | Stage 4 | 0 | 0.0 | 1 | 10.0 | |
| | | Unknown | 2 | 15.4 | 1 | 10.0 | |
| **PMS2** | CRC outcome | CRC diagnosis | 1 | 0.6 | 5 | 1.4 | |
| | | No CRC diagnosis | 179 | 99.4 | 365 | 98.6 | |
| | Stage at diagnosis | Stage 1 | 0 | 0.0 | 3 | 60.0 | 0.500 |
| | | Stage 2 | 0 | 0.0 | 0 | 0.0 | |
| | | Stage 3 | 1 | 100.0 | 0 | 0.0 | |
| | | Stage 4 | 0 | 0.0 | 1 | 20.0 | |
| | | Unknown | 0 | 0.0 | 1 | 20.0 | |
| All genes | CRC outcome | CRC diagnosis | 157 | 10.0 | 177 | 5.6 | |
| | | No CRC diagnosis | 1412 | 90.0 | 2986 | 94.4 | |
| | Stage at diagnosis | Stage 1 | 73 | 46.5 | 62 | 35.0 | 0.040 |
| | | Stage 2 | 29 | 18.5 | 54 | 30.5 | |
| | | Stage 3 | 33 | 21.0 | 37 | 20.9 | |
| | | Stage 4 | 3 | 1.9 | 8 | 4.5 | |
| | | Unknown | 19 | 12.1 | 16 | 9.0 | |

*Denominators for the percentages in colorectal cancer outcome rows are the group totals. Denominators for the percentages in the stage at diagnosis rows are the numbers of colorectal cancers in that specific group. p-values are calculated using the Chi-squared test or Fisher's Exact Test. Tumours are staged at diagnosis. CRC, colorectal cancer; MSVI, mean surveillance interval; all genes, all MMR genes combined.*

#### 3.10. Supplementary table 10: Age-specific five-year annual incidence rates for colorectal cancer for MMR pathogenic variant carriers by mean surveillance interval (≤ 2 years vs > 2 years)

| | Age | **Mean surveillance interval: 2 years** | | | | | | | | | | |
|---|---|---|---|---|---|---|---|---|---|---|---|---|
| | | ≤ 2 years | | | | | > 2 years | | | | | |
| | | No. of CRCs | Obs Years | AS-AIR | Lower CI (95%) | Upper CI (95%) | No. of CRCs | Obs Years | AS-AIR | Lower CI (95%) | Upper CI (95%) | p-value |
| **MLH1** | 25-29 | 4 | 320 | 1252 | 341 | 3205 | 4 | 765 | 523 | 143 | 1339 | 0.357 |
| | 30-34 | 6 | 336 | 1788 | 656 | 3891 | 3 | 909 | 330 | 68 | 964 | 0.089 |
| | 35-39 | 9 | 265 | 3394 | 1552 | 6442 | 8 | 881 | 908 | 392 | 1789 | 0.055 |
| | 40-44 | 16 | 463 | 3455 | 1974 | 5611 | 13 | 1476 | 881 | 468 | 1506 | 0.008 |
| | 50-54 | 5 | 260 | 1926 | 625 | 4494 | 11 | 689 | 1596 | 796 | 2857 | 0.768 |
| | 55-59 | 11 | 230 | 4790 | 2388 | 8571 | 5 | 599 | 835 | 271 | 1949 | 0.016 |
| | 60-64 | 5 | 200 | 2502 | 812 | 5839 | 17 | 483 | 3519 | 2049 | 5635 | 0.518 |
| | 65-69 | 3 | 149 | 2019 | 416 | 5899 | 10 | 299 | 3345 | 1602 | 6152 | 0.465 |
| | 70-74 | 3 | 93 | 3221 | 664 | 9412 | 5 | 194 | 2576 | 837 | 6012 | 0.804 |
| **MSH2** | 25-29 | 5 | 340 | 1469 | 477 | 3428 | 2 | 909 | 220 | 27 | 794 | 0.108 |
| | 30-34 | 8 | 439 | 1824 | 787 | 3593 | 6 | 1109 | 541 | 199 | 1177 | 0.091 |
| | 35-39 | 5 | 433 | 1154 | 375 | 2692 | 7 | 1042 | 672 | 270 | 1384 | 0.463 |
| | 40-44 | 10 | 855 | 1170 | 560 | 2152 | 19 | 2221 | 855 | 515 | 1336 | 0.491 |
| | 50-54 | 13 | 438 | 2970 | 1580 | 5078 | 15 | 1042 | 1440 | 805 | 2375 | 0.118 |
| | 55-59 | 14 | 420 | 3336 | 1822 | 5597 | 15 | 861 | 1742 | 974 | 2873 | 0.139 |
| | 60-64 | 13 | 266 | 4889 | 2600 | 8360 | 13 | 662 | 1964 | 1045 | 3358 | 0.065 |
| | 65-69 | 7 | 242 | 2894 | 1163 | 5962 | 2 | 524 | 382 | 46 | 1379 | 0.048 |
| | 70-74 | 6 | 124 | 4849 | 1779 | 10554 | 7 | 352 | 1986 | 799 | 4093 | 0.231 |
| **MSH6** | 25-29 | | | | | | | | | | | |
| | 30-34 | | | | | | | | | | | |
| | 35-39 | 0 | 148 | 0 | 0 | 2493 | 0 | 341 | 0 | 0 | 1081 | 1.000 |
| | 40-44 | 0 | 369 | 0 | 0 | 1000 | 1 | 804 | 124 | 3 | 693 | 0.688 |
| | 50-54 | 5 | 222 | 2250 | 730 | 5250 | 0 | 490 | 0 | 0 | 754 | 0.054 |
| | 55-59 | 4 | 260 | 1538 | 419 | 3937 | 1 | 583 | 172 | 4 | 956 | 0.142 |
| | 60-64 | 2 | 210 | 955 | 116 | 3448 | 1 | 478 | 209 | 5 | 1166 | 0.408 |
| | 65-69 | 1 | 122 | 821 | 21 | 4573 | 5 | 426 | 1174 | 381 | 2740 | 0.787 |
| | 70-74 | 1 | 108 | 926 | 23 | 5159 | 2 | 315 | 636 | 77 | 2297 | 0.839 |
| **PMS2** | 25-29 | | | | | | | | | | | |
| | 30-34 | | | | | | | | | | | |
| | 35-39 | 0 | 77 | 0 | 0 | 4803 | 0 | 184 | 0 | 0 | 2005 | 1.000 |
| | 40-44 | 0 | 158 | 0 | 0 | 2330 | 1 | 343 | 291 | 7 | 1623 | 0.687 |
| | 50-54 | 0 | 105 | 0 | 0 | 3529 | 0 | 190 | 0 | 0 | 1938 | 1.000 |
| | 55-59 | 1 | 91 | 1100 | 28 | 6131 | 2 | 230 | 871 | 105 | 3146 | 0.895 |
| | 60-64 | 0 | 79 | 0 | 0 | 4690 | 0 | 237 | 0 | 0 | 1555 | 1.000 |
| | 65-69 | 0 | 62 | 0 | 0 | 5964 | 2 | 174 | 1152 | 140 | 4162 | 0.530 |
| | 70-74 | 0 | 41 | 0 | 0 | 8982 | 0 | 131 | 0 | 0 | 2806 | 1.000 |
| **All genes** | 25-29 | 9 | 660 | 1364 | 624 | 2589 | 6 | 1674 | 358 | 132 | 780 | 0.057 |
| | 30-34 | 14 | 774 | 1808 | 988 | 3034 | 9 | 2018 | 446 | 204 | 846 | 0.013 |
| | 35-39 | 14 | 923 | 1516 | 828 | 2544 | 15 | 2448 | 613 | 343 | 1011 | 0.054 |
| | 40-44 | 26 | 1845 | 1409 | 920 | 2065 | 34 | 4844 | 702 | 486 | 981 | 0.026 |
| | 50-54 | 23 | 1024 | 2246 | 1423 | 3370 | 26 | 2411 | 1078 | 704 | 1580 | 0.032 |
| | 55-59 | 30 | 1000 | 2999 | 2023 | 4281 | 23 | 2272 | 1012 | 641 | 1519 | 0.001 |
| | 60-64 | 20 | 754 | 2653 | 1620 | 4097 | 31 | 1860 | 1666 | 1132 | 2365 | 0.162 |
| | 65-69 | 11 | 574 | 1916 | 955 | 3428 | 19 | 1422 | 1336 | 804 | 2086 | 0.415 |
| | 70-74 | 10 | 366 | 2733 | 1308 | 5026 | 14 | 992 | 1411 | 771 | 2367 | 0.200 |

*Only first colorectal cancers are included. Follow up for MSH6 and PMS2 begins at age 35, in line with eligibility for colonoscopic surveillance. 95% confidence intervals are calculated using Byar's method. p-values are the result of a two-sided Z-test. AS-AIR, age-specific annual incidence rate per 100,000; CRC, colorectal cancer.*

#### 3.11. Supplementary table 11: Cumulative risk (in %) of colorectal cancer in MMR pathogenic variant carriers, by mean surveillance interval (≤ 2 years vs > 2 years)

| | | Mean surveillance interval: 2 years | | | | | |
|---|---|---|---|---|---|---|---|
| | | ≤ 2 years | | | > 2 years | | |
| | Age | Cum risk | Lower CI (95%) | Upper CI (95%) | Cum risk | Lower CI (95%) | Upper CI (95%) |
| MLH1 | 25 | 0·0% | 0·0% | 0·0% | 0·8% | 0·0% | 2·2% |
| | 30 | 7·3% | 1·0% | 13·6% | 3·2% | 0·4% | 6·0% |
| | 35 | 15·9% | 6·7% | 25·1% | 4·2% | 1·1% | 7·3% |
| | 40 | 28·0% | 15·8% | 40·2% | 9·0% | 4·6% | 13·5% |
| | 45 | 38·8% | 24·4% | 53·2% | 12·1% | 6·9% | 17·2% |
| | 50 | 50·9% | 34·9% | 66·9% | 17·5% | 11·3% | 23·6% |
| | 55 | 58·4% | 41·6% | 75·2% | 22·8% | 15·7% | 29·8% |
| | 60 | 65·2% | 47·8% | 82·6% | 28·0% | 20·1% | 35·9% |
| | 65 | 69·7% | 51·9% | 87·4% | 39·9% | 30·2% | 49·7% |
| | 70 | 71·9% | 53·9% | 89·9% | 50·0% | 38·6% | 61·4% |
| | 74 | 76·2% | 57·6% | 94·8% | 54·3% | 41·9% | 66·7% |
| MSH2 | 25 | 0·0% | 0·0% | 0·0% | 0·0% | 0·0% | 0·0% |
| | 30 | 6·9% | 0·9% | 12·8% | 1·4% | 0·0% | 2·9% |
| | 35 | 16·8% | 8·4% | 25·2% | 4·5% | 1·7% | 7·2% |
| | 40 | 20·8% | 11·5% | 30·0% | 8·5% | 4·7% | 12·3% |
| | 45 | 26·1% | 16·0% | 36·3% | 12·2% | 7·7% | 16·7% |
| | 50 | 31·8% | 20·8% | 42·7% | 15·7% | 10·7% | 20·7% |
| | 55 | 38·6% | 26·8% | 50·3% | 21·8% | 15·9% | 27·6% |
| | 60 | 51·3% | 38·2% | 64·4% | 27·7% | 21·1% | 34·3% |
| | 65 | 61·5% | 47·3% | 75·7% | 34·3% | 26·8% | 41·9% |
| | 70 | 68·7% | 53·8% | 83·7% | 36·4% | 28·5% | 44·3% |
| | 74 | 72·6% | 57·2% | 88·0% | 41·6% | 32·7% | 50·5% |
| MSH6 | 35 | 0·0% | 0·0% | 0·0% | 0·0% | 0·0% | 0·0% |
| | 40 | 0·0% | 0·0% | 0·0% | 0·0% | 0·0% | 0·0% |
| | 45 | 0·0% | 0·0% | 0·0% | 0·0% | 0·0% | 0·0% |
| | 50 | 2·2% | 0·0% | 6·5% | 1·2% | 0·0% | 3·4% |
| | 55 | 14·0% | 3·9% | 24·1% | 1·2% | 0·0% | 3·4% |
| | 60 | 20·5% | 8·7% | 32·4% | 2·1% | 0·0% | 4·9% |
| | 65 | 20·5% | 8·7% | 32·4% | 5·4% | 0·7% | 10·0% |
| | 70 | 24·3% | 10·5% | 38·0% | 10·1% | 3·6% | 16·6% |
| | 74 | 27·9% | 12·6% | 43·2% | 11·4% | 4·4% | 18·4% |
| PMS2 | 35 | 0·0% | 0·0% | 0·0% | 0·0% | 0·0% | 0·0% |
| | 40 | 0·0% | 0·0% | 0·0% | 0·0% | 0·0% | 0·0% |
| | 45 | 2·9% | 0·0% | 8·4% | 0·0% | 0·0% | 0·0% |
| | 50 | 2·9% | 0·0% | 8·4% | 0·0% | 0·0% | 0·0% |
| | 55 | 2·9% | 0·0% | 8·4% | 4·8% | 0·0% | 13·8% |
| | 60 | 6·6% | 0·0% | 14·0% | 4·8% | 0·0% | 13·8% |
| | 65 | 6·6% | 0·0% | 14·0% | 4·8% | 0·0% | 13·8% |
| | 70 | 13·2% | 1·9% | 24·5% | 4·8% | 0·0% | 13·8% |
| | 74 | 13·2% | 1·9% | 24·5% | 4·8% | 0·0% | 13·8% |
| All genes | 25 | 0·0% | 0·0% | 0·0% | 0·4% | 0·0% | 1·1% |
| | 30 | 7·1% | 2·8% | 11·4% | 2·2% | 0·7% | 3·8% |
| | 35 | 16·1% | 10·0% | 22·2% | 4·3% | 2·2% | 6·3% |
| | 40 | 21·5% | 14·7% | 28·3% | 7·8% | 5·2% | 10·3% |
| | 45 | 26·9% | 19·5% | 34·3% | 10·6% | 7·6% | 13·6% |
| | 50 | 33·4% | 25·5% | 41·4% | 14·1% | 10·7% | 17·4% |
| | 55 | 40·8% | 32·3% | 49·3% | 18·3% | 14·5% | 22·0% |
| | 60 | 49·3% | 40·3% | 58·3% | 22·6% | 18·5% | 26·7% |
| | 65 | 55·0% | 45·6% | 64·4% | 29·2% | 24·5% | 33·9% |
| | 70 | 59·6% | 49·9% | 69·4% | 34·1% | 28·9% | 39·3% |
| | 74 | 63·4% | 53·3% | 73·5% | 37·4% | 31·8% | 42·9% |

*Risk is assumed to be zero below the age of 25 for MLH1 and MSH2 carriers, and below the age of 35 for MSH6 and PMS2 carriers· Cumulative risks are calculated using life table methods, from age-specific annual incidence rates derived from the cohort/analysis groups· Standard errors were calculated using Greenwood's formula, and hence the 95% confidence interval· Cum risk, Cumulative risk; CI, Confidence interval (95%)·*

#### 3.12. Supplementary table 12: Stage-and age-specific five-year annual incidence rates for all MMR pathogenic variant carriers by mean surveillance interval (≤ 2 years vs > 2 years)

| | | **Mean surveillance interval: 2 years** | | | | | | | | | | |
|---|---|---|---|---|---|---|---|---|---|---|---|---|
| | | **≤ 2 years** | | | | | **> 2 years** | | | | | |
| | Five year age band | **No· of CRCs** | **Obs Years** | **AS-AIR** | **Lower CI (95%)** | **Upper CI (95%)** | **No· of CRCs** | **Obs Years** | **AS-AIR** | **Lower CI (95%)** | **Upper CI (95%)** | **p-value** |
| **Stage 1 and 2** | 25-29 | 5 | 660 | 758 | 246 | 1768 | 3 | 1674 | 179 | 37 | 524 | 0·156 |
| | 30-34 | 9 | 774 | 1162 | 531 | 2206 | 4 | 2018 | 198 | 54 | 507 | **0·029** |
| | 35-39 | 7 | 923 | 758 | 305 | 1562 | 10 | 2448 | 408 | 196 | 751 | 0·319 |
| | 40-44 | 14 | 1845 | 759 | 414 | 1273 | 24 | 4844 | 495 | 317 | 737 | 0·280 |
| | 50-54 | 15 | 1024 | 1465 | 819 | 2416 | 17 | 2411 | 705 | 411 | 1129 | 0·089 |
| | 55-59 | 24 | 1000 | 2399 | 1537 | 3570 | 13 | 2272 | 572 | 304 | 978 | **0·001** |
| | 60-64 | 14 | 754 | 1857 | 1014 | 3116 | 19 | 1860 | 1021 | 615 | 1595 | 0·158 |
| | 65-69 | 7 | 574 | 1219 | 490 | 2512 | 17 | 1422 | 1195 | 696 | 1914 | 0·968 |
| | 70-74 | 7 | 366 | 1913 | 769 | 3941 | 9 | 992 | 907 | 415 | 1721 | 0·250 |
| **Stage 3 and 4** | 25-29 | 3 | 660 | 455 | 94 | 1329 | 3 | 1674 | 179 | 37 | 524 | 0·416 |
| | 30-34 | 5 | 774 | 646 | 210 | 1507 | 4 | 2018 | 198 | 54 | 507 | 0·202 |
| | 35-39 | 4 | 923 | 433 | 118 | 1109 | 5 | 2448 | 204 | 66 | 477 | 0·403 |
| | 40-44 | 6 | 1845 | 325 | 119 | 708 | 7 | 4844 | 145 | 58 | 298 | 0·265 |
| | 50-54 | 6 | 1024 | 586 | 215 | 1275 | 8 | 2411 | 332 | 143 | 654 | 0·397 |
| | 55-59 | 4 | 1000 | 400 | 109 | 1024 | 4 | 2272 | 176 | 48 | 451 | 0·380 |
| | 60-64 | 4 | 754 | 531 | 145 | 1358 | 11 | 1860 | 591 | 295 | 1058 | 0·868 |
| | 65-69 | 2 | 574 | 348 | 42 | 1258 | 0 | 1422 | 0 | 0 | 259 | 0·272 |
| | 70-74 | 2 | 366 | 547 | 66 | 1974 | 3 | 992 | 302 | 62 | 883 | 0·645 |

*Only first cancers are included· Tumours are staged at diagnosis· P-values are the result of a two-tailed Z-test· 95% confidence intervals are calculated using Byar's method· AS-AIR, age-specific annual incidence rate pr 100,000 ; CRC, colorectal cancer; LCI, Lower Confidence Interval (95%); MSVI, mean surveillance interval; UCI, Upper Confidence Interval (95%), Years, years of observation·*

#### 3.13. Supplementary table 13: Cumulative risk of CRC-specific death in MMR PV carriers with a mean surveillance interval of ≤ 3 years and > 3 years

| | ≤ 3 years | | | > 3 years | | |
|---|---|---|---|---|---|---|
| **Years of follow up** | **Mortality** | **Lower CI (95%)** | **Upper CI (95%)** | **Mortality** | **Lower CI (95%)** | **Upper CI (95%)** |
| 1 | 0·00% | 0·00% | 0·00% | 0·14% | 0·00% | 0·33% |
| 2 | 0·07% | 0·00% | 0·18% | 0·21% | 0·00% | 0·46% |
| 5 | 0·11% | 0·00% | 0·24% | 0·61% | 0·15% | 1·07% |
| 7 | 0·18% | 0·00% | 0·36% | 1·36% | 0·56% | 2·16% |
| 10 | 0·29% | 0·01% | 0·58% | 1·57% | 0·67% | 2·47% |
| 12 | 0·29% | 0·01% | 0·58% | 2·15% | 0·94% | 3·35% |

*CRC-specific mortality at 1, 2, 5, 7, 10, and 12 years of follow up in the full Lynch syndrome cohort. CRC-specific deaths are defined as any death where a CRC ICD-10 code (C18, C19, C20) appears as a cause code in any section (1A, 1B, 1C, or 2) of the death certificate. Cumulative risk of death is estimated using Kaplan-Meier methods. A total of 22 CRC-specific deaths occurred in the cohort ( 5 in the ≤ 3 years interval group, 17 in the > 3 years group).*

#### 3.14. Supplementary table 14: Associations between mean surveillance interval of ≤ 3 years, sex, age, IMD quintile, and risk of death from colorectal cancer

| | | Hazard Ratio | Lower CI (95%) | Upper CI (95%) | p-value |
|---|---|---|---|---|---|
| Surveillance status | MSVI > 3 years | | | | |
| | MSVI ≤ 3 years | 0·14 | 0·05 | 0·39 | <0·001 |
| Sex | Male | | | | |
| | Female | 0·80 | 0·34 | 1·9 | 0·617 |
| Age at start of follow up | 25-34 | | | | |
| | 35-44 | 1·55 | 0·26 | 9·32 | 0·629 |
| | 45-54 | 4·69 | 0·99 | 22·28 | 0·052 |
| | 55-64 | 3·31 | 0·62 | 17·27 | 0·156 |
| | 65-74 | 7·18 | 1·29 | 40·06 | 0·025 |
| IMD quintile | IMD Q1 - Most deprived | | | | |
| | IMD Q2 | 0·65 | 0·17 | 2·46 | 0·525 |
| | IMD Q3 | 0·83 | 0·26 | 2·76 | 0·761 |
| | IMD Q4 | 0·37 | 0·09 | 1·55 | 0·172 |
| | IMD Q5 - Least deprived | 0·56 | 0·15 | 2·1 | 0·388 |

*Hazard ratios (HR) are estimated using a multivariable Cox Proportional Hazards regression. 4,583 individuals are included in the model, with 149 excluded due to missing data. MMR gene was excluded from the model as no events occurred in MSH6 or PMS2 PV carriers. All included variables met the proportional hazards assumption. Ethnicity, self-reported; IMD, index of multiple deprivation, derived from LSOA of residence; MMR, mismatch repair; MSVI, mean surveillance interval; Sex, person stated gender. P-values are estimated using the Wald Test.*

#### 3.15. Supplementary table 15: Cumulative risk of death of any cause in MMR PV carriers with a mean surveillance interval of ≤ 3 years and > 3 years

| | **≤ 3 years** | | | **> 3 years** | | |
|---|---|---|---|---|---|---|
| **Years of follow up** | **Mortality** | **Lower CI (95%)** | **Upper CI (95%)** | **Mortality** | **Lower CI (95%)** | **Upper CI (95%)** |
| 1 | 0·14% | 0·00% | 0·27% | 1·39% | 0·80% | 1·98% |
| 2 | 0·36% | 0·14% | 0·58% | 1·85% | 1·15% | 2·54% |
| 5 | 1·25% | 0·80% | 1·70% | 3·75% | 2·65% | 4·84% |
| 7 | 2·07% | 1·44% | 2·70% | 5·90% | 4·37% | 7·41% |
| 10 | 3·61% | 2·63% | 4·58% | 7·81% | 5·89% | 9·69% |
| 12 | 4·58% | 3·37% | 5·78% | 9·50% | 7·17% | 11·76% |

*Cumulative risk of death is estimated using Kaplan-Meier methods. A total of 148 deaths occurred in the cohort (68 in the ≤ 2 years interval group, 80 in the > 3 years group).*

#### 3.16. Supplementary table 16: Associations between mean surveillance interval of ≤ 3 years, MMR gene, sex, age, IMD quintile, and risk of death from any cause

| | | **Hazard Ratio** | **Lower CI (95%)** | **Upper CI (95%)** | **p-value** |
|---|---|---|---|---|---|
| Surveillance status | MSVI > 3 years | | | | |
| | MSVI £ 3 years | 0·44 | 0·31 | 0·61 | <0·001 |
| MMR gene | MLH1 | | | | |
| | MSH2 | 1·03 | 0·71 | 1·50 | 0·877 |
| | MSH6 | 0·53 | 0·32 | 0·88 | 0·014 |
| | PMS2 | 0·35 | 0·16 | 0·79 | 0·011 |
| Sex | Male | | | | |
| | Female | 0·70 | 0·50 | 0·97 | 0·032 |
| Age at start of follow up | 25-34 | | | | |
| | 35-44 | 2·58 | 1·04 | 6·41 | 0·042 |
| | 45-54 | 5·79 | 2·53 | 13·28 | <0·001 |
| | 55-64 | 11·43 | 5·10 | 25·63 | <0·001 |
| | 65-74 | 39·24 | 17·59 | 87·52 | <0·001 |
| IMD quintile | IMD Q1 - Most deprived | | | | |
| | IMD Q2 | 0·70 | 0·43 | 1·15 | 0·16 |
| | IMD Q3 | 0·63 | 0·37 | 1·02 | 0·061 |
| | IMD Q4 | 0·57 | 0·35 | 0·94 | 0·028 |
| | IMD Q5 - Least deprived | 0·35 | 0·20 | 0·62 | <0·001 |

*Hazard ratios (HR) are estimated using a multivariable Cox Proportional Hazards regression. 4,583 individuals are included in the model, with 149 excluded due to missing data. All included variables met the proportional hazards assumption. Ethnicity, self-reported; IMD, index of multiple deprivation, derived from LSOA of residence; MMR, mismatch repair; MSVI, mean surveillance interval; Sex, person stated gender. P-values are estimated using the Wald Test.*

#### 3.17. Supplementary table 17: Description of Lynch syndrome cohort for sensitivity analyses and analysis groups

| | | Total | | Mean colonoscopy interval: 2 years | | | | Mean colonoscopy interval: 3 years | | | |
|---|---|---|---|---|---|---|---|---|---|---|---|
| | | | | ≤ 2 years | | > 2 years | | ≤ 3 years | | > 3 years | |
| | | n | % | n | % | n | % | n | % | n | % |
| Total | | 4129 | | 1373 | 33·3 | 2756 | 66·7 | 2832 | 68·6 | 1297 | 31·4 |
| MMR Gene | MLH1 | 1188 | 28·8 | 369 | 26·9 | 819 | 29·7 | 796 | 28·1 | 392 | 30·2 |
| | MSH2 | 1666 | 40·3 | 543 | 39·5 | 1123 | 40·7 | 1159 | 40·9 | 507 | 39·1 |
| | MSH6 | 836 | 20·2 | 315 | 22·9 | 521 | 18·9 | 599 | 21·2 | 237 | 18·3 |
| | PMS2 | 439 | 10·6 | 146 | 10·6 | 293 | 10·6 | 278 | 9·8 | 161 | 12·4 |
| | Unknown | 0 | 0·0 | 0 | 0·0 | 0 | 0·0 | 0 | 0·0 | 0 | 0·0 |
| Sex | Male | 1613 | 39·1 | 559 | 40·7 | 1054 | 38·2 | 1120 | 39·5 | 493 | 38·0 |
| | Female | 2516 | 60·9 | 814 | 59·3 | 1702 | 61·8 | 1712 | 60·5 | 804 | 62·0 |
| | Unknown | 0 | 0·0 | 0 | 0·0 | 0 | 0·0 | 0 | 0·0 | 0 | 0·0 |
| Age at start of follow up period | 25-34 | 1099 | 26·6 | 330 | 24·0 | 769 | 27·9 | 772 | 27·3 | 327 | 25·2 |
| | 35-44 | 1047 | 25·4 | 345 | 25·1 | 702 | 25·5 | 728 | 25·7 | 319 | 24·6 |
| | 45-54 | 912 | 22·1 | 327 | 23·8 | 585 | 21·2 | 653 | 23·1 | 259 | 20·0 |
| | 55-64 | 752 | 18·2 | 263 | 19·2 | 489 | 17·7 | 478 | 16·9 | 274 | 21·1 |
| | 65-74 | 319 | 7·7 | 108 | 7·9 | 211 | 7·7 | 201 | 7·1 | 118 | 9·1 |
| | Unknown | 0 | 0·0 | 0 | 0·0 | 0 | 0·0 | 0 | 0·0 | 0 | 0·0 |
| Ethnicity | White | 3456 | 83·7 | 1205 | 87·8 | 2251 | 81·7 | 2487 | 87·8 | 969 | 74·7 |
| | Asian | 174 | 4·2 | 51 | 3·7 | 123 | 4·5 | 101 | 3·6 | 73 | 5·6 |
| | Black | 18 | 0·4 | 6 | 0·4 | 12 | 0·4 | 13 | 0·5 | 5 | 0·4 |
| | Mixed | 29 | 0·7 | 10 | 0·7 | 19 | 0·7 | 19 | 0·7 | 10 | 0·8 |
| | Other | 83 | 2·0 | 26 | 1·9 | 57 | 2·1 | 53 | 1·9 | 30 | 2·3 |
| | Unknown | 369 | 8·9 | 75 | 5·5 | 294 | 10·7 | 159 | 5·6 | 210 | 16·2 |
| IMD Quintile | Q 1 - most deprived | 647 | 15·7 | 216 | 15·7 | 431 | 15·6 | 428 | 15·1 | 219 | 16·9 |
| | Q 2 | 745 | 18·0 | 254 | 18·5 | 491 | 17·8 | 502 | 17·7 | 243 | 18·7 |
| | Q 3 | 800 | 19·4 | 248 | 18·1 | 552 | 20·0 | 537 | 19·0 | 263 | 20·3 |
| | Q 4 | 909 | 22·0 | 309 | 22·5 | 600 | 21·8 | 642 | 22·7 | 267 | 20·6 |
| | Q 5 - least deprived | 908 | 22·0 | 313 | 22·8 | 595 | 21·6 | 652 | 23·0 | 256 | 19·7 |
| | Unknown | 120 | 2·9 | 33 | 2·4 | 87 | 3·2 | 71 | 2·5 | 49 | 3·8 |

*Includes only individuals with one year of follow up or more. Numbers (n) and percentages (%) are provided for a) the entire cohort (Total), and b) analysis groups as determined by mean surveillance interval. Carriers of PVs in EPCAM (n=23) are included in the MSH2 category. Sex is person stated gender. Ethnicity is self-described. IMD quintile is determined by LSOA of residence at the time of extraction from the Lynch Registry (November 2024).*

#### 3.18. Supplementary table 18: Sensitivity analyses -colorectal cancer diagnoses and stage distribution in a) the whole cohort, and b) analysis groups, for MMR pathogenic variant carriers

| | | | Total | | Mean surveillance interval: 2 years | | | | | Mean surveillance interval: 3 years | | | | |
|---|---|---|---|---|---|---|---|---|---|---|---|---|---|---|
| | | | | | ≤ 2 years | | > 2 years | | | ≤ 3 years | | > 3 years | | |
| | | | n | % | n | % | n | % | p-value | n | % | n | % | p-value |
| MLH1 | CRC | CRC | 118 | 9·9 | 48 | 13·0 | 70 | 8·5 | | 83 | 10·4 | 35 | 8·9 | |
| | | No CRC | 1070 | 90·1 | 321 | 87·0 | 749 | 91·5 | | 713 | 89·6 | 357 | 91·1 | |
| | Stage | Stage 1 | 48 | 40·7 | 25 | 52·1 | 23 | 32·9 | 0·019 | 39 | 47·0 | 9 | 25·7 | 0·019 |
| | | Stage 2 | 32 | 27·1 | 6 | 12·5 | 26 | 37·1 | | 17 | 20·5 | 15 | 42·9 | |
| | | Stage 3 | 22 | 18·6 | 9 | 18·8 | 13 | 18·6 | | 14 | 16·9 | 8 | 22·9 | |
| | | Stage 4 | 4 | 3·4 | 1 | 2·1 | 3 | 4·3 | | 2 | 2·4 | 2 | 5·7 | |
| | | Unk | 12 | 10·2 | 7 | 14·6 | 5 | 7·1 | | 11 | 13·3 | 1 | 2·9 | |
| MSH2 | CRC | CRC | 121 | 7·3 | 53 | 9·8 | 68 | 6·1 | | 90 | 7·8 | 31 | 6·1 | |
| | | No CRC | 1545 | 92·7 | 490 | 90·2 | 1055 | 93·9 | | 1069 | 92·2 | 476 | 93·9 | |
| | Stage | Stage 1 | 53 | 43·8 | 25 | 47·2 | 28 | 41·2 | 0·832 | 41 | 45·6 | 12 | 38·7 | 0·929 |
| | | Stage 2 | 28 | 23·1 | 11 | 20·8 | 17 | 25·0 | | 20 | 22·2 | 8 | 25·8 | |
| | | Stage 3 | 24 | 19·8 | 9 | 17·0 | 15 | 22·1 | | 17 | 18·9 | 7 | 22·6 | |
| | | Stage 4 | 3 | 2·5 | 1 | 1·9 | 2 | 2·9 | | 2 | 2·2 | 1 | 3·2 | |
| | | Unk | 13 | 10·7 | 7 | 13·2 | 6 | 8·8 | | 10 | 11·1 | 3 | 9·7 | |
| MSH6 | CRC | CRC | 12 | 1·4 | 7 | 2·2 | 5 | 1·0 | | 11 | 1·8 | 1 | 0·4 | |
| | | No CRC | 824 | 98·6 | 308 | 97·8 | 516 | 99·0 | | 588 | 98·2 | 236 | 99·6 | |
| | Stage | Stage 1 | 6 | 50·0 | 5 | 71·4 | 1 | 20·0 | 0·098 | 6 | 54·5 | 0 | 0·0 | 0·500 |
| | | Stage 2 | 2 | 16·7 | 0 | 0·0 | 2 | 40·0 | | 1 | 9·1 | 1 | 100 | |
| | | Stage 3 | 2 | 16·7 | 1 | 14·3 | 1 | 20·0 | | 2 | 18·2 | 0 | 0·0 | |
| | | Stage 4 | 1 | 8·3 | 0 | 0·0 | 1 | 20·0 | | 1 | 9·1 | 0 | 0·0 | |
| | | Unk | 1 | 8·3 | 1 | 14·3 | 0 | 0·0 | | 0 | 0·0 | 0 | 0·0 | |
| PMS2 | CRC | CRC | 3 | 0·7 | 0 | 0·0 | 3 | 1·0 | | 0 | 0·0 | 3 | 1·9 | |
| | | No CRC | 436 | 99·3 | 146 | 100·0 | 290 | 99·0 | | 278 | 100·0 | 158 | 98·1 | |
| | Stage | Stage 1 | 2 | 66·7 | 0 | 0·0 | 2 | 66·7 | | 0 | 0·0 | 2 | 66·7 | |
| | | Stage 2 | 0 | 0·0 | 0 | 0·0 | 0 | 0·0 | | 0 | 0·0 | 0 | 0·0 | |
| | | Stage 3 | 0 | 0·0 | 0 | 0·0 | 0 | 0·0 | | 0 | 0·0 | 0 | 0·0 | |
| | | Stage 4 | 1 | 33·3 | 0 | 0·0 | 1 | 33·3 | | 0 | 0·0 | 1 | 33·3 | |
| | | Unk | 0 | 0·0 | 0 | 0·0 | 0 | 0·0 | | 0 | 0·0 | 0 | 0·0 | |
| All genes | CRC | CRC | 254 | 6·2 | 108 | 7·9 | 146 | 5·3 | | 184 | 6·5 | 70 | 5·4 | |
| | | No CRC | 3875 | 93·8 | 1265 | 92·1 | 2610 | 94·7 | | 2648 | 93·5 | 1227 | 94·6 | |
| | Stage | Stage 1 | 109 | 42·9 | 55 | 50·9 | 54 | 37·0 | 0·012 | 86 | 46·7 | 23 | 32·9 | 0·042 |
| | | Stage 2 | 62 | 24·4 | 17 | 15·7 | 45 | 30·8 | | 38 | 20·7 | 24 | 34·3 | |
| | | Stage 3 | 48 | 18·9 | 19 | 17·6 | 29 | 19·9 | | 33 | 17·9 | 15 | 21·4 | |
| | | Stage 4 | 9 | 3·5 | 2 | 1·9 | 7 | 4·8 | | 5 | 2·7 | 4 | 5·7 | |
| | | Unk | 26 | 10·2 | 15 | 13·9 | 11 | 7·5 | | 22 | 12·0 | 4 | 5·7 | |

*Includes only individuals with at least one year of follow up or more. Denominators for the percentages in colorectal cancer outcome rows are the group totals. Denominators for the percentages in the stage at diagnosis rows are the numbers of colorectal cancers in that specific group. p-values are calculated using the Chi-squared test or Fisher's Exact Test. Tumours are staged at diagnosis. CRC, colorectal cancer; all genes, all MMR genes combined*

#### 3.19. Supplementary table 19: Sensitivity analyses - age-specific annual incidence rates by five year age band for colorectal cancer in MMR pathogenic variant carriers

| | Age | Mean surveillance interval: 2 years | | | | | | | | | | | Mean surveillance interval: 3 years | | | | | | | | | | |
|---|---|---|---|---|---|---|---|---|---|---|---|---|---|---|---|---|---|---|---|---|---|---|---|
| | | ≤ 2 years | | | | | > 2 years | | | | | | ≤ 3 years | | | | | > 3 years | | | | | |
| | | No. CRC | Obs Yrs | AS-AIR | LCI (95%) | UCI (95%) | No. CRC | Obs Yrs | AS-AIR | LCI (95%) | UCI (95%) | p-value | No. CRC | Obs Yrs | AS-AIR | LCI (95%) | UCI (95%) | No. CRC | Obs Yrs | AS-AIR | LCI (95%) | UCI (95%) | p-value |
| MLH1 | 25-29 | 3 | 310 | 967 | 199 | 2826 | 3 | 752 | 399 | 82 | 1166 | 0·433 | 3 | 693 | 433 | 89 | 1265 | 3 | 369 | 813 | 168 | 2377 | 0·551 |
| | 30-34 | 4 | 334 | 1198 | 326 | 3067 | 3 | 905 | 331 | 68 | 968 | 0·239 | 5 | 885 | 565 | 184 | 1319 | 2 | 355 | 564 | 68 | 2037 | 0·998 |
| | 35-39 | 7 | 263 | 2661 | 1070 | 5482 | 8 | 877 | 912 | 394 | 1797 | 0·139 | 13 | 819 | 1586 | 844 | 2713 | 2 | 321 | 623 | 75 | 2251 | 0·188 |
| | 40-44 | 11 | 460 | 2394 | 1193 | 4283 | 13 | 1473 | 883 | 470 | 1510 | 0·069 | 19 | 1403 | 1354 | 815 | 2114 | 5 | 529 | 945 | 307 | 2206 | 0·486 |
| | 50-54 | 4 | 258 | 1549 | 422 | 3966 | 9 | 685 | 1314 | 601 | 2493 | 0·818 | 9 | 647 | 1392 | 636 | 2642 | 4 | 297 | 1348 | 367 | 3452 | 0·963 |
| | 55-59 | 9 | 229 | 3930 | 1797 | 7460 | 5 | 596 | 840 | 273 | 1959 | **0·040** | 12 | 525 | 2286 | 1180 | 3993 | 2 | 300 | 668 | 81 | 2412 | 0·083 |
| | 60-64 | 5 | 199 | 2511 | 815 | 5860 | 15 | 481 | 3120 | 1745 | 5146 | 0·695 | 13 | 399 | 3257 | 1732 | 5569 | 7 | 281 | 2494 | 1003 | 5139 | 0·596 |
| | 65-69 | 2 | 147 | 1361 | 165 | 4916 | 9 | 297 | 3032 | 1386 | 5755 | 0·310 | 5 | 263 | 1900 | 617 | 4435 | 6 | 181 | 3320 | 1218 | 7226 | 0·434 |
| | 70-74 | 3 | 93 | 3243 | 669 | 9477 | 5 | 191 | 2622 | 851 | 6119 | 0·813 | 4 | 166 | 2406 | 656 | 6161 | 4 | 117 | 3419 | 932 | 8755 | 0·678 |
| MSH2 | 25-29 | 2 | 334 | 599 | 73 | 2163 | 2 | 897 | 223 | 27 | 805 | 0·509 | 4 | 858 | 466 | 127 | 1194 | 0 | 374 | 0 | 0 | 987 | 0·209 |
| | 30-34 | 4 | 435 | 920 | 251 | 2355 | 4 | 1103 | 363 | 99 | 928 | 0·334 | 7 | 1119 | 626 | 251 | 1289 | 1 | 419 | 239 | 6 | 1329 | 0·367 |
| | 35-39 | 2 | 430 | 465 | 56 | 1680 | 5 | 1035 | 483 | 157 | 1127 | 0·970 | 6 | 1085 | 553 | 203 | 1203 | 1 | 380 | 263 | 7 | 1467 | 0·521 |
| | 40-44 | 8 | 848 | 943 | 407 | 1859 | 13 | 2212 | 588 | 313 | 1005 | 0·386 | 15 | 2233 | 672 | 376 | 1108 | 6 | 828 | 725 | 266 | 1577 | 0·890 |
| | 50-54 | 6 | 431 | 1393 | 511 | 3032 | 13 | 1039 | 1251 | 666 | 2140 | 0·849 | 13 | 1034 | 1258 | 669 | 2151 | 6 | 436 | 1375 | 505 | 2993 | 0·874 |
| | 55-59 | 10 | 414 | 2413 | 1155 | 4438 | 12 | 857 | 1401 | 723 | 2447 | 0·284 | 15 | 866 | 1733 | 969 | 2858 | 7 | 405 | 1726 | 694 | 3557 | 0·994 |
| | 60-64 | 10 | 264 | 3784 | 1812 | 6960 | 10 | 657 | 1523 | 729 | 2801 | 0·110 | 15 | 597 | 2511 | 1404 | 4142 | 5 | 323 | 1546 | 502 | 3607 | 0·361 |
| | 65-69 | 5 | 239 | 2090 | 679 | 4877 | 2 | 522 | 383 | 46 | 1384 | 0·129 | 6 | 497 | 1208 | 443 | 2630 | 1 | 264 | 378 | 10 | 2107 | 0·283 |
| | 70-74 | 6 | 121 | 4958 | 1820 | 10792 | 7 | 348 | 2014 | 810 | 4150 | 0·228 | 9 | 271 | 3315 | 1516 | 6294 | 4 | 197 | 2029 | 553 | 5196 | 0·449 |
| MSH6 | 35-39 | 0 | 140 | 0 | 0 | 2638 | 0 | 329 | 0 | 0 | 1123 | 1·000 | 0 | 303 | 0 | 0 | 1216 | 0 | 165 | 0 | 0 | 2237 | 1·000 |
| | 40-44 | 0 | 360 | 0 | 0 | 1026 | 1 | 794 | 126 | 3 | 702 | 0·691 | 1 | 839 | 119 | 3 | 664 | 0 | 315 | 0 | 0 | 1170 | 0·728 |
| | 50-54 | 4 | 218 | 1831 | 499 | 4689 | 0 | 485 | 0 | 0 | 761 | 0·092 | 4 | 530 | 755 | 206 | 1934 | 0 | 173 | 0 | 0 | 2127 | 0·280 |
| | 55-59 | 1 | 254 | 394 | 10 | 2195 | 0 | 574 | 0 | 0 | 643 | 0·498 | 1 | 596 | 168 | 4 | 934 | 0 | 231 | 0 | 0 | 1597 | 0·722 |
| | 60-64 | 1 | 207 | 482 | 12 | 2688 | 1 | 474 | 211 | 5 | 1174 | 0·715 | 2 | 483 | 414 | 50 | 1496 | 0 | 199 | 0 | 0 | 1854 | 0·490 |
| | 65-69 | 1 | 120 | 836 | 21 | 4655 | 2 | 421 | 475 | 58 | 1716 | 0·774 | 2 | 343 | 583 | 71 | 2107 | 1 | 198 | 506 | 13 | 2817 | 0·930 |
| | 70-74 | 0 | 104 | 0 | 0 | 3540 | 1 | 311 | 322 | 8 | 1793 | 0·750 | 1 | 261 | 383 | 10 | 2132 | 0 | 154 | 0 | 0 | 2403 | 0·640 |
| PMS2 | 35-39 | 0 | 72 | 0 | 0 | 5095 | 0 | 178 | 0 | 0 | 2072 | 1·000 | 0 | 139 | 0 | 0 | 2647 | 0 | 111 | 0 | 0 | 3320 | 1·000 |
| | 40-44 | 0 | 152 | 0 | 0 | 2419 | 1 | 336 | 297 | 8 | 1658 | 0·690 | 0 | 313 | 0 | 0 | 1179 | 1 | 176 | 569 | 14 | 3168 | 0·508 |
| | 50-54 | 0 | 102 | 0 | 0 | 3634 | 0 | 189 | 0 | 0 | 1955 | 1·000 | 0 | 207 | 0 | 0 | 1778 | 0 | 83 | 0 | 0 | 4459 | 1·000 |
| | 55-59 | 0 | 88 | 0 | 0 | 4192 | 1 | 224 | 446 | 11 | 2484 | 0·720 | 0 | 209 | 0 | 0 | 1768 | 1 | 104 | 965 | 24 | 5377 | 0·502 |
| | 60-64 | 0 | 77 | 0 | 0 | 4790 | 0 | 234 | 0 | 0 | 1575 | 1·000 | 0 | 196 | 0 | 0 | 1882 | 0 | 115 | 0 | 0 | 3200 | 1·000 |
| | 65-69 | 0 | 60 | 0 | 0 | 6187 | 1 | 171 | 584 | 15 | 3256 | 0·743 | 0 | 151 | 0 | 0 | 2440 | 1 | 80 | 1257 | 32 | 7005 | 0·505 |
| | 70-74 | 0 | 39 | 0 | 0 | 9429 | 0 | 125 | 0 | 0 | 2948 | 1·000 | 0 | 92 | 0 | 0 | 3997 | 0 | 72 | 0 | 0 | 5125 | 1·000 |
| All genes | 25-29 | 5 | 644 | 776 | 252 | 1811 | 5 | 1649 | 303 | 98 | 708 | 0·268 | 7 | 1551 | 451 | 181 | 930 | 3 | 742 | 404 | 83 | 1181 | 0·889 |
| | 30-34 | 8 | 769 | 1041 | 449 | 2050 | 7 | 2009 | 348 | 140 | 718 | 0·111 | 12 | 2004 | 599 | 309 | 1046 | 3 | 774 | 388 | 80 | 1133 | 0·520 |
| | 35-39 | 9 | 905 | 994 | 455 | 1887 | 13 | 2419 | 537 | 286 | 919 | 0·253 | 19 | 2348 | 809 | 487 | 1264 | 3 | 977 | 307 | 63 | 898 | 0·084 |
| | 40-44 | 19 | 1820 | 1044 | 628 | 1630 | 28 | 4816 | 581 | 386 | 840 | 0·099 | 35 | 4787 | 731 | 509 | 1017 | 12 | 1848 | 649 | 335 | 1134 | 0·735 |
| | 50-54 | 14 | 1009 | 1388 | 758 | 2328 | 22 | 2398 | 918 | 575 | 1389 | 0·298 | 26 | 2417 | 1076 | 702 | 1576 | 10 | 989 | 1011 | 484 | 1859 | 0·877 |
| | 55-59 | 20 | 985 | 2030 | 1239 | 3135 | 18 | 2250 | 800 | 474 | 1264 | **0·019** | 28 | 2196 | 1275 | 847 | 1843 | 10 | 1040 | 962 | 460 | 1769 | 0·455 |
| | 60-64 | 16 | 748 | 2140 | 1222 | 3475 | 26 | 1846 | 1408 | 920 | 2064 | 0·256 | 30 | 1675 | 1791 | 1208 | 2556 | 12 | 918 | 1307 | 674 | 2283 | 0·366 |
| | 65-69 | 8 | 566 | 1415 | 611 | 2787 | 14 | 1411 | 992 | 542 | 1665 | 0·499 | 13 | 1254 | 1037 | 552 | 1773 | 9 | 723 | 1246 | 570 | 2365 | 0·706 |
| | 70-74 | 9 | 357 | 2522 | 1153 | 4788 | 13 | 974 | 1335 | 710 | 2282 | 0·240 | 14 | 791 | 1769 | 966 | 2968 | 8 | 540 | 1483 | 640 | 2921 | 0.711 |

*Included only individuals with one year of follow up or more. Incidence is per 100,000. Only first colorectal cancers are included. Follow up for MSH6 and PMS2 begins at age 35, in line with eligibility for colonoscopic surveillance. 95% confidence intervals are calculated using Byar's method. AS-AIR, age-specific annual incidence rate per 100,000 ; CRC, colorectal cancer; LCI, lower confidence interval; UCI, upper confidence interval; Yrs, years.*

### 3.20. Supplementary table 20: Sensitivity analyses - cumulative risk colorectal cancer from age 25-74 in MMR pathogenic variant carriers

| | | Mean surveillance interval: 2 years | | | | | | Mean surveillance interval: 3 years | | | | | |
|---|---|---|---|---|---|---|---|---|---|---|---|---|---|
| | | ≤ 2 years | | | > 2 years | | | ≤ 3 years | | | > 3 years | | |
| | Age | Cum risk | LCI (95%) | UCI (95%) | Cum risk | LCI (95%) | UCI (95%) | Cum risk | LCI (95%) | UCI (95%) | Cum risk | LCI (95%) | UCI (95%) |
| MLH1 | 25 | 0·0 | 0·0 | 0·0 | 0·0 | 0·0 | 0·0 | 0·0 | 0·0 | 0·0 | 0·0 | 0·0 | 0·0 |
| | 30 | 2·5 | 0·1 | 4·8 | 4·6 | 0·0 | 9·7 | 2·2 | 0·0 | 4·6 | 5·2 | 0·2 | 10·3 |
| | 35 | 3·5 | 0·7 | 6·3 | 12·1 | 3·9 | 20·3 | 5·4 | 1·9 | 9·0 | 6·6 | 0·9 | 12·3 |
| | 40 | 8·4 | 4·2 | 12·6 | 21·6 | 10·7 | 32·6 | 12·2 | 7·1 | 17·4 | 11·0 | 3·5 | 18·4 |
| | 45 | 11·5 | 6·5 | 16·5 | 30·2 | 17·0 | 43·4 | 17·7 | 11·4 | 24·0 | 12·8 | 4·5 | 21·0 |
| | 50 | 16·9 | 10·8 | 23·0 | 38·9 | 24·1 | 53·7 | 23·6 | 16·4 | 30·8 | 20·4 | 9·9 | 31·0 |
| | 55 | 21·1 | 14·3 | 27·9 | 47·1 | 31·2 | 63·0 | 30·7 | 22·5 | 38·9 | 23·1 | 12·0 | 34·3 |
| | 60 | 26·5 | 18·7 | 34·2 | 54·8 | 38·0 | 71·6 | 37·9 | 28·7 | 47·1 | 28·1 | 16·0 | 40·2 |
| | 65 | 37·4 | 27·9 | 46·9 | 59·4 | 42·0 | 76·7 | 47·5 | 36·9 | 58·0 | 35·8 | 22·3 | 49·3 |
| | 70 | 47·1 | 35·8 | 58·4 | 62·4 | 44·6 | 80·2 | 51·7 | 40·4 | 63·0 | 48·2 | 32·1 | 64·2 |
| | 74 | 51·9 | 39·4 | 64·4 | 68·2 | 49·4 | 87·1 | 57·6 | 44·9 | 70·2 | 53·5 | 35·9 | 71·0 |
| MSH2 | 25 | 0·0 | 0·0 | 0·0 | 0·0 | 0·0 | 0·0 | 0·0 | 0·0 | 0·0 | 0·0 | 0·0 | 0·0 |
| | 30 | 1·4 | 0·0 | 2·9 | 2·6 | 0·0 | 6·2 | 2·4 | 0·3 | 4·5 | 0·0 | 0·0 | 0·0 |
| | 35 | 3·6 | 1·1 | 6·1 | 8·0 | 2·1 | 13·8 | 6·2 | 3·0 | 9·5 | 1·2 | 0·0 | 3·4 |
| | 40 | 5·9 | 2·7 | 9·1 | 10·2 | 3·6 | 16·8 | 8·4 | 4·7 | 12·2 | 3·7 | 0·0 | 7·8 |
| | 45 | 8·5 | 4·7 | 12·2 | 15·3 | 7·4 | 23·2 | 11·6 | 7·3 | 16·0 | 7·2 | 1·5 | 12·8 |
| | 50 | 11·7 | 7·3 | 16·1 | 20·0 | 11·1 | 28·9 | 15·1 | 10·2 | 20·0 | 11·5 | 4·5 | 18·5 |
| | 55 | 17·3 | 12·0 | 22·6 | 22·8 | 13·4 | 32·3 | 19·3 | 13·8 | 24·8 | 17·5 | 9·0 | 25·9 |
| | 60 | 21·6 | 15·6 | 27·6 | 35·6 | 24·0 | 47·1 | 27·4 | 20·7 | 34·1 | 22·5 | 13·0 | 32·0 |
| | 65 | 27·8 | 20·7 | 34·8 | 44·9 | 31·8 | 58·1 | 35·2 | 27·4 | 43·1 | 28·7 | 17·9 | 39·6 |
| | 70 | 30·1 | 22·6 | 37·5 | 54·5 | 39·7 | 69·2 | 41·1 | 32·3 | 49·9 | 31·7 | 20·1 | 43·3 |
| | 74 | 35·9 | 27·1 | 44·6 | 60·2 | 44·4 | 76·0 | 47·5 | 37·5 | 57·4 | 36·7 | 23·9 | 49·5 |
| MSH6 | 35 | 0·0 | 0·0 | 0·0 | 0·0 | 0·0 | 0·0 | 0·0 | 0·0 | 0·0 | 0·0 | 0·0 | 0·0 |
| | 40 | 0·0 | 0·0 | 0·0 | 0·0 | 0·0 | 0·0 | 0·0 | 0·0 | 0·0 | 0·0 | 0·0 | 0·0 |
| | 45 | 0·0 | 0·0 | 0·0 | 0·0 | 0·0 | 0·0 | 0·0 | 0·0 | 0·0 | 0·0 | 0·0 | 0·0 |
| | 50 | 2·3 | 0·0 | 6·6 | 1·2 | 0·0 | 3·5 | 2·2 | 0·0 | 5·2 | 0·0 | 0·0 | 0·0 |
| | 55 | 8·6 | 0·4 | 16·7 | 1·2 | 0·0 | 3·5 | 4·8 | 0·6 | 9·0 | 0·0 | 0·0 | 0·0 |
| | 60 | 12·1 | 2·6 | 21·5 | 1·2 | 0·0 | 3·5 | 6·5 | 1·7 | 11·3 | 0·0 | 0·0 | 0·0 |
| | 65 | 12·1 | 2·6 | 21·5 | 2·3 | 0·0 | 5·5 | 7·6 | 2·3 | 12·8 | 0·0 | 0·0 | 0·0 |
| | 70 | 16·2 | 4·0 | 28·4 | 4·8 | 0·2 | 9·5 | 10·6 | 4·0 | 17·3 | 2·6 | 0·0 | 7·6 |
| | 74 | 16·2 | 4·0 | 28·4 | 6·2 | 0·9 | 11·6 | 12·3 | 4·9 | 19·7 | 2·6 | 0·0 | 7·6 |
| PMS2 | 35 | 0·0 | 0·0 | 0·0 | 0·0 | 0·0 | 0·0 | 0·0 | 0·0 | 0·0 | 0·0 | 0·0 | 0·0 |
| | 40 | 0·0 | 0·0 | 0·0 | 0·0 | 0·0 | 0·0 | 0·0 | 0·0 | 0·0 | 0·0 | 0·0 | 0·0 |
| | 45 | 0·0 | 0·0 | 0·0 | 3·0 | 0·0 | 8·8 | 0·0 | 0·0 | 0·0 | 5·1 | 0·0 | 14·4 |
| | 50 | 0·0 | 0·0 | 0·0 | 3·0 | 0·0 | 8·8 | 0·0 | 0·0 | 0·0 | 5·1 | 0·0 | 14·4 |
| | 55 | 0·0 | 0·0 | 0·0 | 3·0 | 0·0 | 8·8 | 0·0 | 0·0 | 0·0 | 5·1 | 0·0 | 14·4 |
| | 60 | 0·0 | 0·0 | 0·0 | 4·9 | 0·0 | 11·8 | 0·0 | 0·0 | 0·0 | 8·9 | 0·0 | 20·8 |
| | 65 | 0·0 | 0·0 | 0·0 | 4·9 | 0·0 | 11·8 | 0·0 | 0·0 | 0·0 | 8·9 | 0·0 | 20·8 |
| | 70 | 0·0 | 0·0 | 0·0 | 8·4 | 0·0 | 17·8 | 0·0 | 0·0 | 0·0 | 17·2 | 0·0 | 36·1 |
| | 74 | 0·0 | 0·0 | 0·0 | 8·4 | 0·0 | 17·8 | 0·0 | 0·0 | 0·0 | 17·2 | 0·0 | 36·1 |
| All genes | 25 | 0·0 | 0·0 | 0·0 | 0·0 | 0·0 | 0·0 | 0·0 | 0·0 | 0·0 | 0·0 | 0·0 | 0·0 |
| | 30 | 1·9 | 0·5 | 3·2 | 3·7 | 0·5 | 6·8 | 2·3 | 0·7 | 3·9 | 2·5 | 0·1 | 5·0 |
| | 35 | 3·5 | 1·6 | 5·3 | 9·6 | 4·8 | 14·5 | 5·7 | 3·4 | 8·1 | 3·8 | 0·8 | 6·8 |
| | 40 | 6·2 | 3·9 | 8·5 | 13·6 | 8·0 | 19·1 | 9·1 | 6·3 | 12·0 | 6·2 | 2·5 | 9·9 |
| | 45 | 8·6 | 5·9 | 11·2 | 18·3 | 12·0 | 24·5 | 12·3 | 9·1 | 15·5 | 8·8 | 4·5 | 13·1 |
| | 50 | 11·9 | 8·9 | 15·0 | 23·3 | 16·4 | 30·1 | 16·1 | 12·5 | 19·6 | 13·1 | 8·0 | 18·2 |
| | 55 | 15·6 | 12·1 | 19·0 | 28·0 | 20·7 | 35·4 | 20·3 | 16·4 | 24·3 | 16·5 | 10·9 | 22·1 |
| | 60 | 18·9 | 15·2 | 22·7 | 36·1 | 28·1 | 44·1 | 25·9 | 21·5 | 30·3 | 20·4 | 14·3 | 26·5 |
| | 65 | 24·6 | 20·2 | 29·0 | 41·3 | 32·8 | 49·8 | 31·8 | 26·8 | 36·7 | 25·3 | 18·6 | 32·1 |
| | 70 | 28·8 | 24·0 | 33·7 | 47·0 | 37·8 | 56·2 | 35·9 | 30·5 | 41·2 | 31·3 | 23·7 | 38·8 |
| | 74 | 32·4 | 27·1 | 37·8 | 51·4 | 41·6 | 61·2 | 40·2 | 34·3 | 46·1 | 34·5 | 26·4 | 42·6 |

*Includes only individuals with at least one year of follow up or more. Risk is assumed to be zero below the age of 25 for MLH1 and MSH2 carriers, and below the age of 35 for MSH6 and PMS2 carriers. Cumulative risks are calculated using life table methods, from age-specific annual incidence rates derived from the cohort/analysis groups. Standard errors were calculated using Greenwood's formula, and hence the 95% confidence interval. Cum risk, Cumulative risk; L/UCI, Lower/Upper Confidence interval (95%).*

### 3.21. Supplementary table 21: Sensitivity analyses - stage- and age-specific annual incidence rates for all MMR pathogenic variant carriers combined

| | Age | Mean surveillance interval: 2 years | | | | | | | | | | | Mean surveillance interval: 3 years | | | | | | | | | | |
|---|---|---|---|---|---|---|---|---|---|---|---|---|---|---|---|---|---|---|---|---|---|---|---|
| | | ≤ 2 years | | | | | > 2 years | | | | | | ≤ 3 years | | | | | > 3 years | | | | | |
| | | No· CRC | Obs Yrs | AS-AIR | LCI | UCI | No· CRC | Obs Yrs | AS-AIR | LCI | UCI | p-value | No· CRC | Obs Yrs | AS-AIR | LCI | UCI | No· CRC | Obs Yrs | AS-AIR | LCI | UCI | p-value |
| Stage 1 and 2 | 25-29 | 3 | 644 | 466 | 96 | 1361 | 2 | 1649 | 121 | 15 | 438 | 0.312 | 4 | 1551 | 258 | 70 | 660 | 1 | 742 | 135 | 3 | 750 | 0.612 |
| | 30-34 | 6 | 769 | 780 | 286 | 1699 | 3 | 2009 | 149 | 31 | 436 | 0.092 | 7 | 2004 | 349 | 140 | 720 | 2 | 774 | 258 | 31 | 934 | 0.740 |
| | 35-39 | 4 | 905 | 442 | 120 | 1131 | 9 | 2419 | 372 | 170 | 706 | 0.811 | 12 | 2348 | 511 | 264 | 893 | 1 | 977 | 102 | 3 | 570 | 0.059 |
| | 40-44 | 10 | 1820 | 549 | 263 | 1011 | 21 | 4816 | 436 | 270 | 667 | 0.599 | 24 | 4787 | 501 | 321 | 746 | 7 | 1848 | 379 | 152 | 780 | 0.526 |
| | 50-54 | 9 | 1009 | 892 | 408 | 1693 | 15 | 2398 | 626 | 350 | 1032 | 0.473 | 17 | 2417 | 703 | 409 | 1126 | 7 | 989 | 708 | 285 | 1458 | 0.990 |
| | 55-59 | 16 | 985 | 1624 | 928 | 2637 | 13 | 2250 | 578 | 307 | 988 | 0.026 | 20 | 2196 | 911 | 556 | 1407 | 9 | 1040 | 866 | 396 | 1643 | 0.906 |
| | 60-64 | 12 | 748 | 1605 | 828 | 2804 | 15 | 1846 | 813 | 454 | 1340 | 0.151 | 20 | 1675 | 1194 | 729 | 1844 | 7 | 918 | 762 | 306 | 1571 | 0.316 |
| | 65-69 | 6 | 566 | 1061 | 389 | 2309 | 13 | 1411 | 921 | 490 | 1576 | 0.804 | 11 | 1254 | 877 | 437 | 1570 | 8 | 723 | 1107 | 478 | 2182 | 0.659 |
| | 70-74 | 6 | 357 | 1681 | 617 | 3660 | 8 | 974 | 821 | 355 | 1618 | 0.306 | 9 | 791 | 1137 | 520 | 2159 | 5 | 540 | 927 | 301 | 2162 | 0.739 |
| Stage 3 and 4 | 25-29 | 1 | 644 | 155 | 4 | 865 | 3 | 1649 | 182 | 38 | 532 | 0.916 | 2 | 1551 | 129 | 16 | 466 | 2 | 742 | 269 | 33 | 973 | 0.598 |
| | 30-34 | 2 | 769 | 260 | 32 | 940 | 3 | 2009 | 149 | 31 | 436 | 0.662 | 4 | 2004 | 200 | 54 | 511 | 1 | 774 | 129 | 3 | 720 | 0.746 |
| | 35-39 | 2 | 905 | 221 | 27 | 798 | 4 | 2419 | 165 | 45 | 423 | 0.800 | 4 | 2348 | 170 | 46 | 436 | 2 | 977 | 205 | 25 | 740 | 0.869 |
| | 40-44 | 4 | 1820 | 220 | 60 | 563 | 4 | 4816 | 83 | 23 | 213 | 0.319 | 4 | 4787 | 84 | 23 | 214 | 4 | 1848 | 216 | 59 | 554 | 0.326 |
| | 50-54 | 4 | 1009 | 396 | 108 | 1015 | 6 | 2398 | 250 | 92 | 545 | 0.572 | 7 | 2417 | 290 | 116 | 597 | 3 | 989 | 303 | 63 | 886 | 0.955 |
| | 55-59 | 2 | 985 | 203 | 25 | 733 | 3 | 2250 | 133 | 27 | 390 | 0.731 | 5 | 2196 | 228 | 74 | 531 | 0 | 1040 | 0 | 0 | 355 | 0.123 |
| | 60-64 | 3 | 748 | 401 | 83 | 1173 | 10 | 1846 | 542 | 259 | 996 | 0.676 | 8 | 1675 | 477 | 206 | 941 | 5 | 918 | 544 | 177 | 1271 | 0.842 |
| | 65-69 | 1 | 566 | 177 | 4 | 985 | 0 | 1411 | 0 | 0 | 261 | 0.495 | 1 | 1254 | 80 | 2 | 444 | 0 | 723 | 0 | 0 | 511 | 0.644 |
| | 70-74 | 2 | 357 | 560 | 68 | 2025 | 3 | 974 | 308 | 64 | 900 | 0.642 | 3 | 791 | 379 | 78 | 1108 | 2 | 540 | 371 | 45 | 1339 | 0.984 |

*Includes only individuals with at least one year of follow up or more. Only first colorectal cancers are included. Follow up for MSH6 and PMS2 begins at age 35, in line with eligibility for colonoscopic surveillance. 95% confidence intervals are calculated using Byar's method. Tumours are staged at diagnosis. AS-AIR, age-specific annual incidence rate per 100,000 ; CRC, colorectal cancer; Obs, observation; LCI, lower confidence interval (95%); UCI, upper confidence interval (95%); Yrs, years..*
